# Does early childhood BCG vaccination improve survival into adulthood in a population with a low tuberculosis prevalence? Quasi-experimental evidence on non-specific effects from 39 Swedish birth cohorts

**DOI:** 10.1101/2023.02.16.23286013

**Authors:** Michaela Theilmann, Pascal Geldsetzer, Till Bärnighausen, Nikkil Sudharsanan

## Abstract

The Bacillus Calmette–Guérin (BCG) vaccine for tuberculosis (TB) is widely used globally. Many high-income countries discontinued nationwide vaccination policies as the TB prevalence decreased. However, there is continued interest in whether the general childhood immunity boost conferred by the BCG vaccination impacts adult health and mortality in low-TB contexts (known as non-specific effects) and whether BCG vaccination should be continued as a population policy. While recent studies found evidence of an association between BCG vaccination and later-life survival, it is unclear whether these associations are causal or driven by unobserved characteristics of those who chose to voluntarily vaccinate. We use the abrupt discontinuation of mandatory BCG vaccination in Sweden in 1975 as a natural experiment to estimate the causal non-specific effect of the BCG vaccine on long-term cohort survival. Applying two complementary study designs, we find no evidence that survival to age 30 was affected by the discontinuation of childhood BCG vaccination. The results are consistent in the male and female subpopulations and are robust to several sensitivity and falsification tests. Overall, despite interest and prior correlational studies suggesting large non-specific effects, we do not find any population-level evidence for a non-specific effect of the BCG vaccine discontinuation on long-term survival in Sweden.

## Introduction

Tuberculosis (TB) was a major cause of mortality in Europe in the 18^th^ century and is estimated to have caused 1,000 deaths per 100,000 people (Murray, 2001). The Bacillus Calmette–Guérin (BCG) vaccine was developed in the 1910s to address the global TB burden and was administered to the first child in 1921 (Dara et al., 2014). BCG vaccination was then scaled up widely in the 1940s across Europe. It became the main government strategy for containing TB and many high-income countries (HIC) introduced national policies making BCG vaccination compulsory for children (Zwerling et al., 2011). As the incidence of TB declined to negligible levels, several HICs discontinued compulsory vaccinations. However, there has been continued and significant scientific interest in the *non-specific* effects of the BCG vaccine and whether it increases long-term survival beyond its effects on TB.

The proposed mechanism behind non-specific BCG effects is that early BCG vaccination improves general childhood immunity, which may confer a lasting health impact as individuals age (Cirovic et al., 2020; Dockrell & Smith, 2017). This hypothesis is consistent with life course theories in demography in population health that link early life conditions and events with later life morbidity and mortality (Case & Paxson, 2010; Cutler & Miller, 2005; Elo & Preston, 1992; Helgertz & Bengtsson, 2019; Masters, 2018; McEnry & Palloni, 2010; Palloni & Souza, 2013; van den Berg, Doblhammer-Reiter, & Christensen, 2011; Wen & Gu, 2011; Zhao, Hessel, Simon Thomas, & Beckfield, 2021). Despite the growing scientific interest over the past two decades, especially among laboratory scientists studying cellular-level immune response to the vaccine, it is still inconclusive whether the early childhood improvement in immune response created by the BCG vaccine has a causal impact on health in later life (Dockrell & Smith, 2017; Goodridge et al., 2016).

There exists a small body of literature analysing the relationship between the BCG vaccine and morbidity and mortality among adults (Giamarellos-Bourboulis et al., 2020; Kölmel et al., 2005; Lankes, Fought, Evens, Weisenburger, & Chiu, 2009; Pfahlberg et al., 2002; Rieckmann; Rieckmann, Villumsen, Jensen, et al., 2017; Rieckmann, Villumsen, Sørup, et al., 2017; Villumsen et al., 2009; Villumsen et al., 2013; Wardhana, Sultana, Mandang, & Jim, 2011). Indeed, a recent study suggested that children vaccinated with the BCG vaccine in Denmark had an 42% lower hazard of mortality in adulthood (Rieckmann, Villumsen, Sørup, et al., 2017). If this association was in fact evidence of a causal relationship, these results would have significant population policy implications and would suggest that re-introducing BCG vaccination in national vaccination plans, even in the absence of TB, may be a cost-effective and easy-to-implement way of improving long-term survival. However, these studies are either based on small non-representative samples, limited to specific health outcomes rather than an overall measure like all-cause mortality, and most fundamentally are based on study designs that suffer from confounding. Thus, it is unclear whether the association between BCG and later-life survival is due to the causal effect of improved childhood immunity from vaccination or reflects the unobserved characteristics of the families and children that opted for the vaccine. To our knowledge, there is no causal evidence on the non-specific effects of the vaccine on long-term survival at the population level.

In this paper, we address these three key limitations. First, rather than rely on small or potentially non-representative samples, we use population data on births and deaths in Sweden for birth cohorts born between 1950 and 1988. Second, rather than focus on specific health conditions, we examine the effects of BCG vaccination on cohort survival to age 30 based on all-cause mortality. Lastly, we employ two causal inference study designs – the Regression Discontinuity Design (RDD) and the multi-period Difference in Differences (DiD) model – to address the potential issues of confounding found in prior studies. In contrast to prior studies, we do not find that the BCG vaccine had non-specific effects on survival to age 30 in Sweden. This result is consistent across analysis approaches and robust to several sensitivity analyses and alternate specifications. While BCG vaccination in the absence of TB might still be important for certain specific subpopulations, our results do not provide evidence in support of broad population policies to re-introduce BCG vaccination in HICs with a low burden of TB.

## Background

### Epidemiology of Tuberculosis

TB is an infectious disease caused by the mycobacterium tuberculosis bacterium that mostly affects the lungs but can also spread to other organs or the bones (Raviglione Mario C., 2018). TB is highly fatal if untreated but can be treated with a six-month antibiotic drug regimen. It is transmitted through the air (e.g. from talking or coughing) and is mainly observed among adults, males, and those that are immunocompromised (e.g. living with HIV), living with diabetes, undernourished, and those that use alcohol or tobacco (Lönnroth, Jaramillo, Williams, Dye, & Raviglione, 2009).

The mycobacterium tuberculosis has been around for thousands of years but tuberculosis prevalence increased to an epidemic level only in the 18^th^ century, reaching an estimated death rate of 1,000 per 100,000 inhabitants in 1800 (Barberis, Bragazzi, Galluzzo, & Martini, 2017; Chisholm, Trauer, Curnoe, & Tanaka, 2016; Hershkovitz et al., 2015; Murray, 2001). This dramatic increase in TB mortality is thought to be related to increasing urbanization and the close agglomeration of individuals in cities as well as poor living standards (Lönnroth et al., 2009). In the 19^th^ century, the TB prevalence declined in high-income countries (Wilson, 1990) While TB remains the second highest cause of death from infectious diseases globally, after COVID-19, in 2020, this burden is concentrated in low and middle-income countries, with high-income countries only accounting for 2% of all recorded TB cases (World Health Organization, 2021).

### Development and Introduction of the BCG Vaccine in Europe

The BCG vaccine is a live vaccine and was introduced in the 1920s (Dara et al., 2014). Since then it has become one of the most widely used vaccines globally (Dara et al., 2014; Muhoza et al., 2021). Several strains of the BCG vaccine emerged from the original strain isolated by Albert Calmette and Camille Guérin in 1908 (Dara et al., 2014). There is conflicting evidence on the effectiveness of the vaccine and to date, it is poorly understood how the BCG vaccine affects the immune system and why there is such a wide variation in the body’s immunological response (Dockrell & Smith, 2017). While evidence is mixed, it is generally believed that the BCG vaccine only protects against the development of active TB but not infection itself, and that the vaccine has the greatest benefits when administered after birth rather than to adolescents or adults (World Health Organization; Abubakar et al., 2013). The BCG vaccine is safe for healthy children but poses a threat to immunodeficient children, who can develop a disseminated BCG infection (World Health Organization, 2018). This fact is important for deciding which children should be given the BCG vaccination, a point which we will return to later on when discussing potential issues of confounding in prior research.

In Europe, the BCG vaccine was widely adopted in the 1940s (Dara et al., 2014). In the large majority of countries it was administered right after birth (Zwerling et al., 2011). While some countries, such as Sweden and Norway, made the BCG vaccination compulsory, other countries adopted policies that targeted high-risk groups only, such as healthcare workers or children of parents born in high-incidence countries. In the decades after the introduction of the BCG vaccine, the TB incidence continued to decrease dramatically in Western Europe and other high-income countries (Daniel, 2006). This decline lead most of the countries where vaccination was compulsory to discontinue their policies (Böttiger, Romanus, Verdier, & Boman, 1982). In Sweden, for example, TB mortality decreased from around 22 per 100,000 in 1950 to less than 4 per 100,000 in 1975 (Committee on Government Operations, United States Congress Senate; Sakari Härö, 1994). In Norway, mortality attributable to TB was 29 per 100,000 people in 1950 and decreased to 2 per 100,000 in 1975 (Central Bureau of Statistics Norway, 1961).^1^ Importantly, it remains unclear how much the BCG vaccine contributed to the decline in TB prevalence, or whether this decline was mainly due to improved living standards and nutrition (Wilson, 1990).

In Sweden, the country we study here, compulsory vaccination of neonates was introduced in 1940 and then reversed in April 1975 because of the persistently low TB prevalence (The Public Health Agency of Sweden, 2020; Bergström, Mäkinen, & Romanus, 2001; Zwerling et al., 2011). Thus, children born in 1976 were the first birth cohort in which no newborn received the routine BCG vaccine. This shift was abrupt: vaccine coverage dropped from 95% to 2% in the years following its discontinuation (Romanus, 2006). Coverage then gradually increased to 13.1% in the cohort born in 1989 and covered at-risk groups such as children born to parents from high TB incidence countries (Bergström et al., 2001). In Norway, BCG vaccinations became compulsory in 1947 for students leaving elementary school, usually at age 13 (Waaler, Galtung, & Mordal, 1971). Mass vaccination campaigns started in 1947 (Tverdak & Funnemark, 1988). However, unlike Sweden, mandatory vaccination in Norway remained until 2005, when the policy changed to a voluntary scheme, and again in 2009, when the vaccination recommendation was limited to at risk groups only (Norwegian Institute of Public Health, 2008; Norwegian Institute of Public Health, 2016). BCG vaccine coverage in Norway increased from 88.6% among adolescents aged 13 in 1959 to 92.2% in 1973 and was above 97% since 1975 (Norwegian Institute of Public Health; Tverdak & Funnemark, 1988).

### Non-Specific Effects of the BCG Vaccine

Non-specific effects are health effects of a vaccine that are unrelated to the targeted disease. The first non-specific effects were reported on the vaccinia vaccine against smallpox, which also seemed to protect against measles and other infectious diseases (Mayr, 2004). These findings led to the assumption that other live vaccines, such as the BCG vaccine, might have positive non-specific effects (Higgins et al., 2016). It is not entirely clear through which channels the BCG vaccine could provide protection against other diseases. There is evidence that the BCG vaccine causes an adaption of the innate, or unspecific, immunity, which is the fraction of the immune system humans have from birth on and is, thus, not induced in response to specific diseases through infection or vaccination (Roberts, Alberts, Bruce, Johnson, Alexander, Lewis, Raff, & Walter, 2015). This trained immunity, i.e. the change in the innate immunity after BCG vaccination, is one of the potential channels for the BCG vaccine related non-specific effects (Moorlag, Arts, van Crevel, & Netea, 2019). This physiological mechanism is consistent with a broader body of evidence from demography and population health which finds a robust link between early childhood health and changes to the immune system and later life (Bonanni, 1999; Drummond, Chevat, & Lothgren, 2007; Karafillakis, Hassounah, & Atchison, 2015; Mercer, 1985; Michiels, Govaerts, Remmen, Vermeire, & Coenen, 2011). Today, the BCG vaccine is widely applied as a treatment for bladder cancer and research on its effectiveness in the treatment of other cancers is ongoing (Lamm & Morales, 2021).

### Existing evidence of non-specific BCG effects

There is a small number of studies, most of them observational, estimating non-specific effects of the BCG vaccine on child survival (Biering-Sørensen et al., 2018; Breiman et al., 2004; Castro, Pardo-Seco, & Martinón-Torres, 2015; Elguero, Simondon, Vaugelade, Marra, & Simondon, 2005; Garly et al., 2003; Lehmann, Vail, Firth, Klerk, & Alpers, 2005; Nankabirwa, Tumwine, Mugaba, Tylleskär, & Sommerfelt, 2015; Roth et al., 2005; Schaltz-Buchholzer, Aaby, et al., 2021; Schaltz-Buchholzer, Kjær Sørensen, Benn, & Aaby, 2021; Steenhuis et al., 2008; Thysen et al., 2020; Vaugelade, Pinchinat, Guiella, Elguero, & Simondon, 2004). The majority of these studies was conducted in low- and middle-income countries and suggest a positive association between BCG vaccine and child survival. Evidence on the vaccine’s non-specific effect on adult morbidity and mortality is also limited (Giamarellos-Bourboulis et al., 2020; Kölmel et al., 2005; Lankes et al., 2009; Pfahlberg et al., 2002; Rieckmann; Rieckmann, Villumsen, Jensen, et al., 2017; Rieckmann, Villumsen, Sørup, et al., 2017; Villumsen et al., 2009; Villumsen et al., 2013; Wardhana et al., 2011). All studies but one are based on small non-representative samples or are limited to specific health outcomes (Rieckmann, Villumsen, Sørup, et al., 2017). Most importantly, all studies except for one clinical trial do not sufficiently account for potential confounding and can thus only estimate associations but not establish causal relationships (Giamarellos-Bourboulis et al., 2020).

For example, Pfahlberg et al. (2002) study the relationship between BCG vaccination and malignant melanoma among 1,230 individuals from eleven health centers in seven European countries between 1994 and 1997. Using a case-control design, the authors compare those that did and did not receive the BCG vaccine and conclude that the BCG vaccine might reduce the risk of malignant melanoma. In contrast, Riekmann et al. (2019) link data on 5,090 adults from the Copenhagen School Health Records to the Danish Cancer Registry and find no association between BCG vaccination and malignant melanoma. Focusing on lymphoma and leukaemia, Villumsen et al. (2009) also use data from the Copenhagen School Health Records and find that individuals who had received the BCG vaccine had half the risk of developing lymphoma compared to those who had not. However, there was no difference in the risk of developing leukaemia.

Two observational studies analyse the association between BCG vaccination and adult survival. Kölmel et al. (2005) analysed the association of the vaccinia and BCG vaccines with survival among individuals with malignant melanoma in six European countries and Israel. The authors compared participants with the vaccine to those without and found no evidence of an association between the BCG vaccine and survival among individuals with malignant melanoma. The only study analysing the relationship between the BCG vaccination and long-term survival used the Copenhagen School Health Records Register used as part of the prior mentioned papers. Rieckmann et al. (2017) linked these data to the Civil Registration System and the Danish Register of Causes of Death databases, focusing on death by natural causes (excluding tuberculosis or smallpox). Comparing those that did receive the BCG vaccine to those that did not, they found that vaccinated individuals had approximately half the risk of dying from natural causes than unvaccinated individuals.

While some of these studies suggest that the BCG vaccine has non-specific effects on morbidity and mortality, none of the study designs can establish a causal relationship^2^ because they do not sufficiently account for confounding due to factors such as the healthy vaccinee bias (Schaltz-Buchholzer, Kjær Sørensen, et al., 2021). The healthy vaccinee bias occurs if healthy children, and thus children with a higher overall probability of survival, are more likely to receive the vaccine than unhealthy children. The decision to not administer the vaccine can be based on concerns around side effects or because these parents have a lower health service uptake in general. Comparing vaccinated to unvaccinated individuals without considering systematic differences in characteristics that are correlated with the decision to vaccinate can result in an over- or underestimation of the vaccine’s true non-specific effects. We address this knowledge gap by applying quasi-experimental methods that take advantage of the sudden discontinuation of compulsory vaccination to estimate the causal effect of BCG vaccination on survival into adulthood.

## Data and methods

### Data

We screened all available information on the BCG Atlas to identify countries that would be eligible for inclusion in our study (Zwerling et al., 2011). First, we identified all countries which had a change in vaccination policy, moving from compulsory to optional vaccination as potential natural experiments. Next, we screened the documentation provided on each of these countries in the Human Mortality Database (HMD) as of January 19, 2022 to identify which of these countries have available data for all years in our study period (University of California, Berkeley, and Max Planck Institute for Demographic Research). We identified six potential countries with a change in vaccination policy that also had data available in the HMD (Austria, Denmark, Finland, France, Great Britain, Norway, Sweden). We chose Sweden as most suitable for the following reasons: (1) there was a sharp discontinuation of compulsory vaccinations in 1975 and BCG vaccine coverage dropped from 95% before 1975 to 2% in the years following the discontinuation (Romanus, 2006); and (2) Sweden was the first country to abolish a nationwide policy and discontinue the BCG vaccine. Thus, Sweden allows for the maximum observation period in terms of survival into adulthood.

One of our study designs, the multi-period DiD, additionally requires selecting a control country that followed a similar survival trajectory prior to the discontinuation of BCG vaccination in Sweden but that did not discontinue vaccines at the same or similar time as Sweden. Based on this criterion, we selected Norway as the most suitable counterfactual country for the DiD analyses.^3^ For both Sweden and Norway, we accessed data on the number of births and age-specific death rates for each birth cohort born from 1846 to 1988 from the Human Mortality Database. To minimize confounding from the aftermath of World War II, we only included birth cohorts born in 1950 or later. Thus, our observation period covers cohorts born from 1950 to 1988, which allows us to study the impact of BCG vaccination on the cohort probability of survival to age 30 (for those born in the final 1988 birth cohort, we have data on their survival up to the year 2018, when cohort members turned 30).

Unfortunately, we were not able to use Swedish register data, which is commonly used in demographic analyses of the Swedish population, (Barclay & Kolk, 2018; Bucher-Koenen, Farbmacher, Guber, & Vikström, 2020; Cook, Fletcher, & Forgues, 2019; Helgertz & Bengtsson, 2019; Lazuka, 2019; Oudin Åström, Edvinsson, Hondula, Rocklöv, & Schumann, 2016) for the following two reasons: (1) our analysis requires survival data for cohorts born several years before the BCG discontinuation in 1975, but the Demographic Data Base only includes data from 1973 and later. (2) It is not currently possible to link the Demographic Data Base to BCG vaccination data.

### Data preparation

Our outcome of interest is the probability of survival to age 30 in each birth cohort by country. Constructing this outcome requires information on the number of individuals that survived to age 30 in each cohort. The HMD data include cohort age-specific death rates but not the total number of deaths that occurred in a birth cohort by age. We, therefore, had to convert the age-specific death rates to death counts. We did this through the following procedure. First, we converted the cohort age-specific mortality rates to the age-specific probabilities of survival using the standard life-table approach (Preston, Heuveline, & Guillot, 2001). Second, we generated a dataset with the number of observations equal to the number of births in a cohort. We then generated a variable for “alive at age 30” and used the cohort life table probability of survival from birth to age 30 to determine what share of this initial birth cohort survived to age 30, and correspondingly assigned values to the “alive at age 30” variable. For example, if among the 115,414 births for the 1950 birth cohort, the cohort-life table probability of survival to age 30 was 90%, we would assign values of 1 for “alive at age 30” to (0.9 x 111,414 =) 100,273 observations. This procedure was repeated for each birth cohort, resulting in a final dataset of 4,152,211 observations, which equals the total number of births in Sweden from 1950 to 1988. We repeated this approach to generate individual-level datasets for Norway as well as separately for males and females in each country.

### Identification strategies

We used two complementary quasi-experimental designs to assess the effect of the sharp BCG discontinuation in Sweden on cohort survival to age 30: the regression discontinuity design (RDD) and the multi-period difference in differences (DiD) approach. The RDD estimates the causal effect of an intervention or policy in situations where exposure to the intervention/policy is based on whether individuals fall on one side or another of some type of cut-off. This cut-off point is usually referred to as the “threshold” and the continuous variable on which the cut off is based is known as the “running variable.” For example, individuals may be eligible for free health care (the policy) based on whether their income (the running variable) is below the poverty line (the threshold). In such circumstances, the RDD design estimates the causal effect of the policy under the assumption that in the absence of the treatment, the relationship between the running variable and an outcome would have evolved continuously. Heuristically, this can be expressed that those just below the threshold are, on average, identical in all characteristics to those just above the threshold. The only difference being that those above the threshold receive the treatment (in our study, this would be not receiving BCG vaccination). Importantly, the RDD estimates the local average treatment effect (LATE) at the threshold value.

For our analysis, we followed the continuity-based RDD approach described by Cattaneo et al. (2019). The first assumption of an RDD is that the probability of being treated changes discontinuously at the specified threshold. In our analysis, the running variable is year of birth and the threshold is 1976, the birth year of the first cohort for which the BCG vaccine was not mandatory anymore. There are no yearly data on BCG coverage for the years around the threshold. However, Romanus et al. (2006) report that BCG vaccine coverage dropped from 95% to 2% in the five years following the discontinuation, which shows that the probability of receiving the BCG vaccine decreased sharply at the threshold. A second assumption is that the outcome would be continuous at the threshold in the absence of the treatment. In other words, there was no substantial change in the probability of survival to age 30 induced by an event other than the BCG vaccination discontinuation. To our knowledge, there have been no major shifts in policies nor national-level events, such as natural disasters, which could have affected the probability of survival of the cohorts born just before the discontinuation differently to those born just after. Finally, for the RDD approach to be valid, individuals should not systematically sort around the threshold with the aim to obtain or avoid the treatment. We assess this risk to be negligible in the context of our study as we consider it unlikely that parents accurately anticipated and based their family planning on the future BCG policy change. This is supported by the data, which shows no clustering of births neither above nor below the threshold (Figure S 1).

We followed the current best practice for RDD estimation (Cattaneo et al., 2019). This includes using a local linear model (to prevent overfitting the data), including triangular Kernel weights (to give more weight to observations closer to the threshold), and a data-driven approach for identifying the bandwidth around the threshold (to remove arbitrary bandwidth selection and potential manipulation of the bandwidth size by researchers) (Imbens & Kalyanaraman, 2012). For inference, we used robust bias-corrected standard errors and confidence intervals (Cattaneo et al., 2019). Our estimation model is:

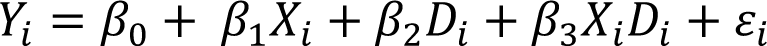

Here *Y_i_* is the probability of survival to age 30 for new born *i, X_i_* is a continuous measure of year of birth (the running variable), and *D_i_* is a binary indicator that takes the value 1 if a birth cohort was born after the discontinuation of mandatory BCG vaccination (Cunningham, 2021). β_3_ is the causal effect of the discontinuation of the BCG vaccine on survival to age 30 for the cohort born in the threshold year, i.e. 1976.

While the RDD estimates the effect of the BCG discontinuation for the cohort born in the threshold year, it could be the case that the effect only emerges after several birth cohorts (for example if it took a few birth cohorts to fully discontinue the vaccine). Although it is unlikely, it could also be the case that another shock unrelated to BCG discontinuation occurred over the same period of the discontinuation. To investigate the effect of BCG vaccination in the case of these two scenarios, we conducted a multi-period DiD model (sometimes also referred to as leads and lags model) with Norway as the comparison country.

The multi-period DiD estimates the effect of the vaccine discontinuation by comparing the probability of survival in Sweden and Norway in each birth cohort accounting for temporal trends, i.e. the general positive trend in the probability of survival, as well as the baseline difference between the two countries. The central identifying assumption of the multi-period DiD is that in the absence of BCG discontinuation, the cohort probability of survival to age 30 in Sweden would follow the same trend as the cohort probability of survival to age 30 in Norway (the so-called parallel trends assumption). This assumption is commonly assessed by comparing the trends in the outcome variable in the pre-treatment period. While we formally assess this assumption later, the visual evidence presented in Figure 1 shows that among several potential comparison countries, the cohort mortality trends in Norway most closely mirrored those in Sweden. By estimating the effect of BCG discontinuation on survival for multiple birth cohorts born after the discontinuation – and not just the 1976 birth cohort as in the RDD - this approach reveals whether the effect wanes or intensifies over time or whether there is a lag in the effect and only cohorts born some years after the discontinuation are affected.

**Figure 1:**
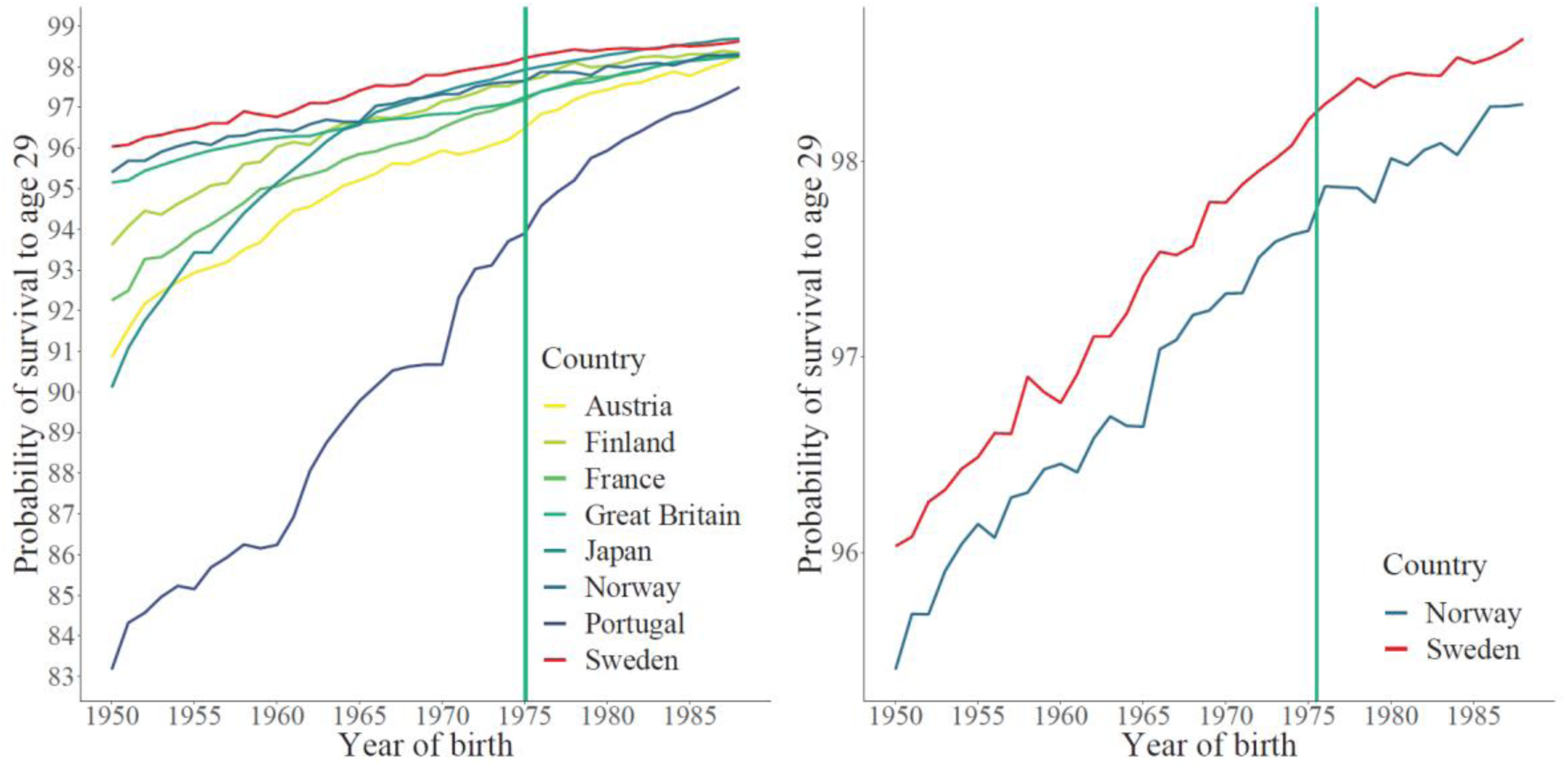
Probability of survival to age 29 in Sweden and potential control countries. Note: We plotted the probability of survival to age 29, rather than 30, because data needed to construct survival to age 30 were not available for France and Great Britain. The vertical line divides the birth cohorts into those born before the vaccine discontinuation (1975 and earlier) and those born after.

The multi-period DiD regression model is set up as follows:

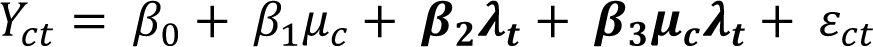

with *c* indexing countries and *t* indexing years (Cunningham, 2021). μ*_c_* is a country fixed effect which accounts for the baseline difference in the probability of survival between the treatment (Sweden) and control (Norway) countries. λ*_t_* is a vector of year fixed effects which account for the time trend common to both countries (1975, the year of the BCG vaccine discontinuation, is the reference year). μ*_t_* λ_t_ is a vector of the interaction between the country and year dummies and can be interpreted as the treatment effect for each of the birth cohorts. Importantly, the coefficients on the country-year interaction terms for cohorts born before 1975 provides a test of the parallel trends assumption (these coefficients should be close to 0). Lastly, rather than estimate treatment effects for each birth cohort born after the BCG discontinuation, we can also estimate a common treatment effect for all birth cohorts by interacting the country dummy with a post dummy (=1 for all cohorts born in or after the year of discontinuation) and including year fixed effects as individual dummy variables (Angrist & Pischke, 2008):

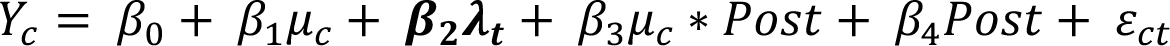

## Results

The final data include one observation per person born from 1950 to 1988. For Sweden, this includes 4,152,211 people in total (2,137,830 men and 2,014,381 women), ranging from 91,780 in the cohort born in 1983 to 123,354 in 1966. In Norway, 2,326,387 people were born in this period (1,196,500 men and 1,129,887 women), ranging from 49,937 in 1983 to 67,746 in 1969. Figure 2 shows the probability of survival to age 30 in Sweden for birth cohorts born between 1950 to 1988. Survival to age 30 increased from 95.9% in the 1950 birth cohort to 98.6% in the 1988 birth cohort. Across all years, the probability of survival was higher among women than men. For example, in 1975 the probability of survival to age 30 was 98.6% among women and 97.7% among men. Despite this difference in the absolute level, survival in the female and male subpopulations followed a similar trend. Visually, we observe no evidence of an abrupt change in the probability of survival in the whole population when comparing the cohort born in the year of the BCG vaccine discontinuation (98.2% in 1975) and the first cohort for whom the vaccine was optional (98.2% in 1976). This smooth trend around 1975 is also present when separately looking at the male and female subpopulations.

**Figure 2:**
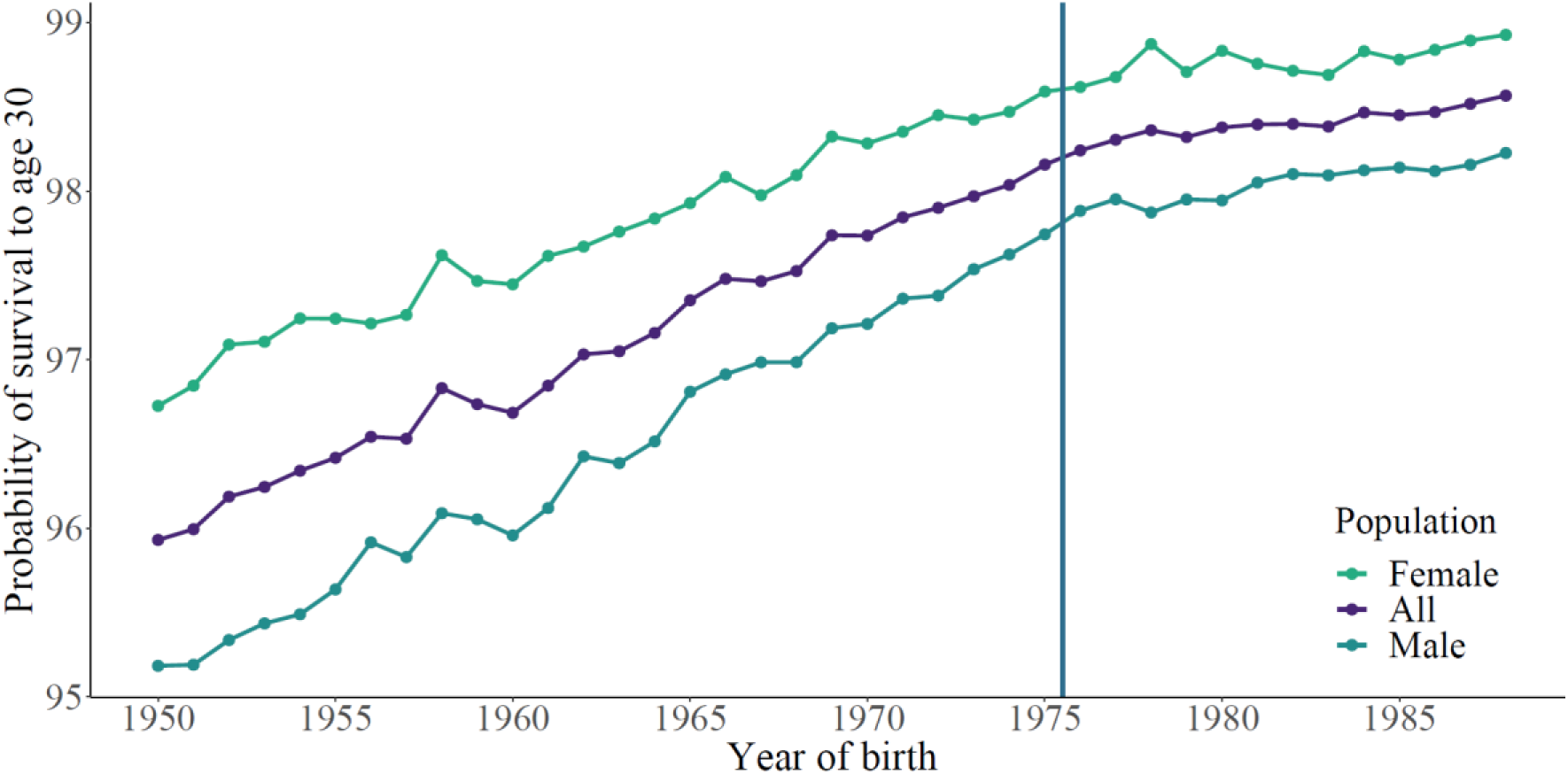
Cohort probability of survival from birth to age 30 in Sweden. Note: The vertical line divides the birth cohorts into those in or born before the year of the vaccine discontinuation (1975 and earlier) and those born after.

### Regression Discontinuity results

The RDD analysis confirms the results of the visual inspection: we do not find evidence of an effect of the BCG vaccination discontinuation on long-term survival in the birth cohort born in 1976. We estimate a precise null effect (0.00 percentage points (pp) (95% CI -0.20 – 0.17)) on the probability of survival to age 30 at the point of BCG vaccination discontinuation (Table 1, Figure 3). As with the visual results, this finding was not driven by just males or females. We also find no evidence that BCG vaccine discontinuation affected the probability of survival to age 30 for among the male (0.05pp (−0.17 - 0.25)) or female (0.11pp (−0.46 – 0.15)) subpopulations (Table 1, Figures S 2 and S 3).

**Table 1:** Regression Discontinuity Design estimation results for the effect of BCG vaccination discontinuation on the cohort probability of survival from birth to age 30 in Sweden.

|  | Total |  | Men |  | Women |  |
| --- | --- | --- | --- | --- | --- | --- |
|  | below<br>cut-off | above<br>cut-off | below<br>cut-off | above<br>cut-off | below<br>cut-off | above<br>cut-off |
| Sample size | 323,169 | 383,905 | 339,754 | 343,064 | 103,683 | 139,722 |
| Bandwidth | 3.95 | 3.95 | 6.09 | 6.09 | 2.87 | 2.87 |
| Local average treatment effect (percentage points) |  |  |  |  |  |  |
| Coefficient | 0.00 |  | 0.05 |  | -0.11 |  |
| 95% CI | (-0.20 - 0.17) |  | (-0.17 - 0.25) |  | (-0.46 - 0.15) |  |
| p-value | 0.86 |  | 0.70 |  | 0.33 |  |

*Note: We estimated the Regression Discontinuity Design model by fitting a local linear trend including triangular Kernel weights, a data-driven bandwidth selection, and a regularization term. The derivation of the optimal bandwidth can result in a decimal. In line with Cattaneo et al. (2019), we used the floor of the decimal. The table displays the local average treatment effect for the birth cohort of 1976 and confidence intervals resulting from robust bias-corrected standard errors.*

**Figure 3:**
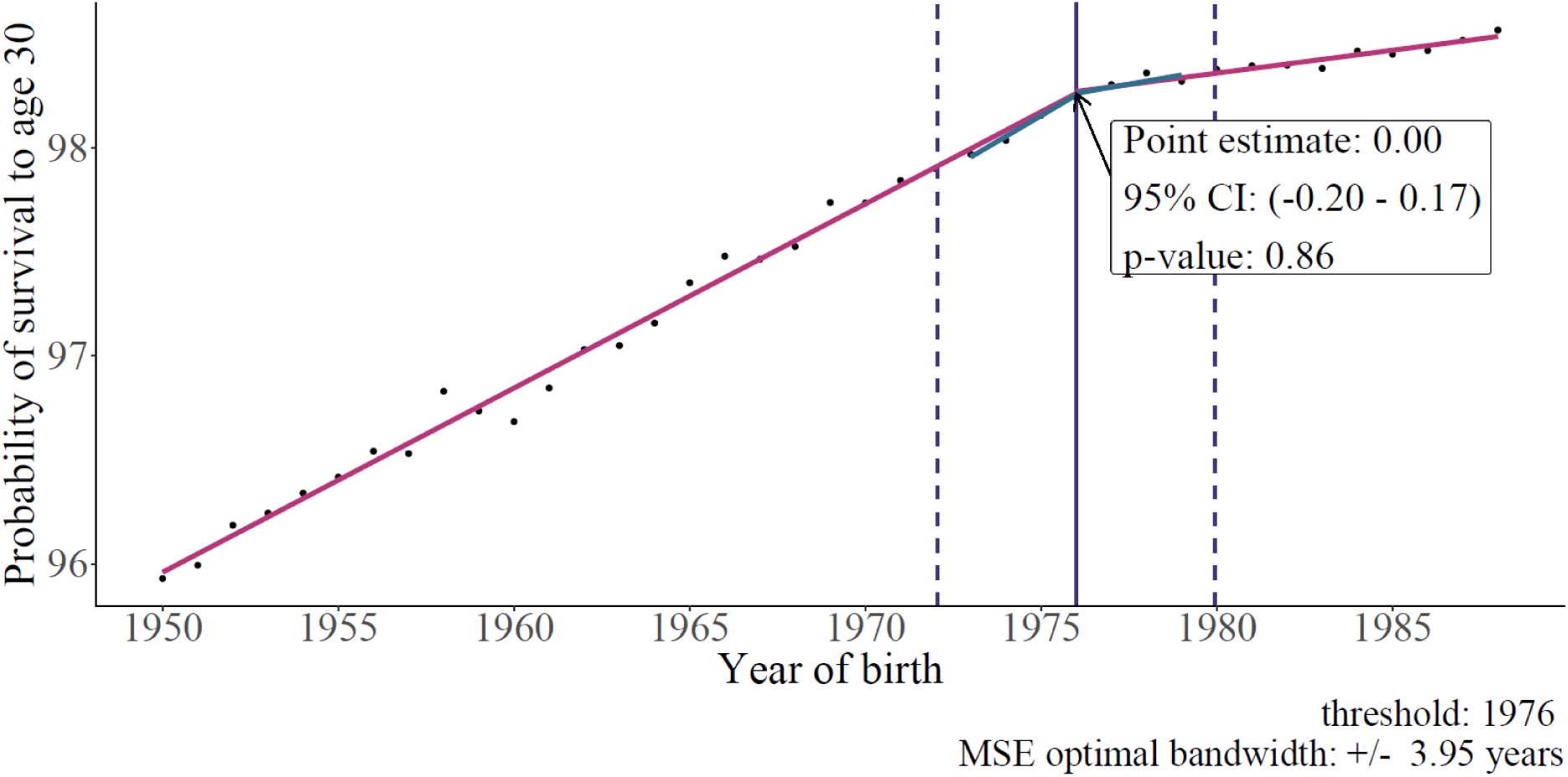
Non-specific effects of the BCG vaccine discontinuation on the cohort probability of survival from birth to age 30 in Sweden. *Note: Pink lines represent linear trends fitted separately for cohorts born before and after vaccine discontinuation; blue lines are local linear trends fitted within the bandwidth and separately for cohorts born before and after vaccine discontinuation; dashed lines represent the bandwidth determined by the data; purple vertical line represents the year 1976, year of birth of the cohort from whom the local average treatment effect is estimated*.

### Multi-period difference in differences results

While the RDD estimates potential non-specific effects for the cohort born in the year of the vaccine discontinuation, the DiD allows us to generate estimates for each birth cohort and, thus, yields the opportunity to detect lagged effects. The event-study plot in Figure 4 shows the difference in survival between birth cohorts born in Norway and Sweden for each cohort born before and after the BCG discontinuation. The coefficients are shown net of an overall level difference in survival between the two countries and a time trend. In support of the parallel trends assumption, we find that most point estimates in the pre-discontinuation period are not statistically different from zero (exact values are shown in Table S 1). This suggests that prior to the discontinuation of BCG vaccination in Sweden, both countries were following a parallel trend in cohort survival to age 30. The coefficients after the vertical line in 1975 capture any effect of BCG discontinuation on survival in Sweden. Similar to our RDD results, we find no evidence of differences in cohort survival to age 30 in the post-BCG-discontinuation period between individuals born in Sweden and Norway across most birth cohorts. For Swedish cohorts born after 1983, we see some evidence of lower survival to age 30; however, these differences are for cohorts born nearly 10 years after the discontinuation of BCG. For this reason, we consider it more likely that the negative point estimates for younger birth cohorts were caused by other changes at the population level that differed between Norway and Sweden at that time rather than being caused by the discontinuation of the BCG vaccine.

**Figure 4:**
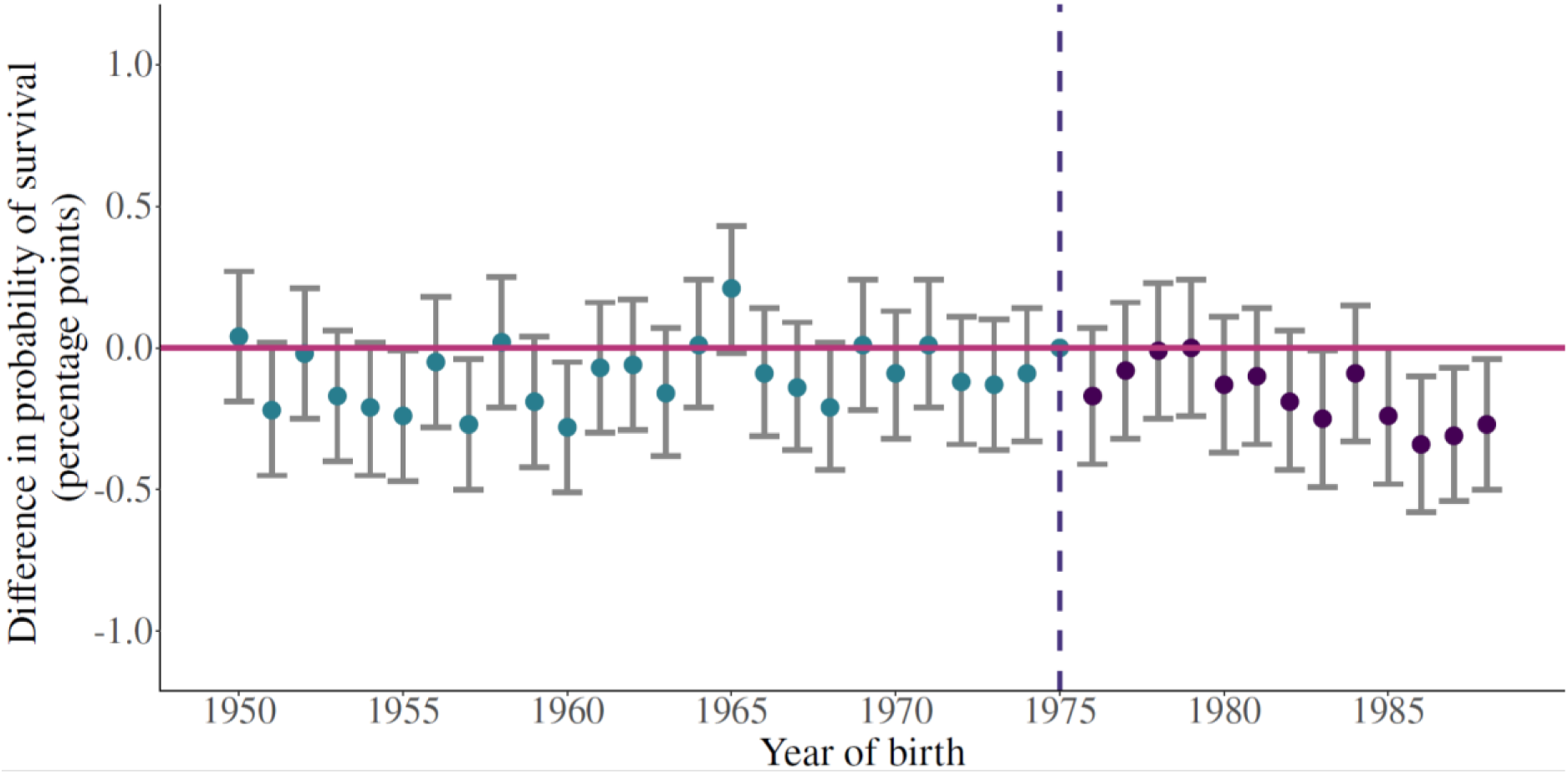
Event study plot displaying multi-period difference in differences results. Note: Coefficients (with 95% confidence intervals) display the difference in the probability of survival between cohorts born in the respective year in Sweden and Norway net the secular time trend and fixed countries differences. The dashed vertical line marks the reference group, which is the cohort born in 1975.

By splitting the results by birth cohort, the event-study results may result in underpowered estimates of the effect of BCG discontinuation. When pooling all data from the pre- and post-discontinuation periods, respectively, we find evidence of a very small negative effect of the BCG vaccine on the probability of survival to age 30 by -0.06pp (−0.11 – 0.00) (Table 2). While these results are statistically precise due to our large sample size (over six million observations), the effects we estimate are extremely small in magnitude at less than a 10th of a percentage point and thus of minimal policy significance. Further, based on the event-study plot, this minimal effect is likely driven by the lower survival among the 1983+ birth cohorts.

**Table 2:** Pooled Difference in Difference estimates

|  |  | Total | Men | Women |
| --- | --- | --- | --- | --- |
| all | country*post | -0.06 | -0.11 | 0.00 |
| observations |  | (-0.11 - 0.00) | (-0.19 - -0.02) | (-0.07 - 0.07) |
|  |  | *0.04* | *0.02* | *0.91* |
| observations | country*post | 0.05 | -0.05 | 0.06 |
| within MSE |  | (-0.06 - 0.16) | (-0.18 - 0.07) | (-0.1 - 0.23) |
| optimal |  | *0.39* | *0.40* | *0.44* |
| bandwidth |  |  |  |  |

*Note: We pooled data from all pre-discontinuation and post-discontinuation years respectively. Regression models include year dummies. The table displays 95% confidence intervals in parenthesis and p-values in italic. The MSE optimal bandwidth was six for the full population, six for the male population, and two for the female population. The cut-off year was 1975.*

We find a similar set of conclusions when examining males and females separately. Among both the male and female population, there was no difference in the probability of survival for the cohorts born before the BCG vaccine discontinuation, indicating support for the parallel trend assumption. In the female subpopulation, there was also no difference in the probability of survival between Sweden and Norway after the vaccine discontinuation (net of the time and level trends). This visual finding is confirmed in the pooled analysis (0.00pp (−0.07 - 0.07)). In the male subpopulation, the results also suggest that there were no non-specific effects of the BCG vaccine on survival. However, as observed in the total population, cohorts born after 1983 in Sweden had a lower probability of survival in Sweden than in Norway. Furthermore, pooling all pre- and post-discontinuation birth cohorts reveals that the probability of survival decreased by 0.11pp (−0.19 – -0.02) among men.

It is likely that this result is driven by the youngest birth cohorts, which represent approximately one third of the post-discontinuation birth cohorts, but were born nearly 10 years after the vaccine discontinuation and thus the validity of the DiD assumption that any difference was due to the discontinuation alone for this group is questionable. To test this, we restricted our sample to the cohorts born within the MSE optional bandwidth calculated in the RDD analysis, which was +/-six years in the male sample (Table 2). The results show that for the birth cohorts born within the MSE optimal bandwidth, there was no difference in the probability of survival between the total, male, and female populations in Sweden and Norway (Table 2).

### Sensitivity analyses

We conducted several sensitivity analyses to test the robustness of our results. First, it could be the case that the rate of change of survival across cohorts depends on the absolute level of survival, and that this functional form affects our estimates. To investigate this, we plotted the log probability of survival and found that flattening of the cohort survival trends was not driven by the fact that the probability of survival approached 100% (Figure S 6). Second, the RDD estimates whether there was a sudden change in the level of the probability of survival after the BCG vaccine discontinuation. It could, however, also be the case that the discontinuation slowed down the rate of change in the increase of the probability of survival to age 30. To investigate this, we applied a Regression Kink Design (RKD). The RKD relies on the same assumptions as the RDD but is applied when there is not a discontinuity in the probability of being treated but in its first derivative, i.e. in the slope of the relationship between the treatment and the assignment variable. In the context of our analysis, this would be the case if the probability of receiving the vaccine was still high in 1975, the year of the discontinuation, and only gradually decreased in the following years. Another potential scenario could be that the probability of being vaccinated already declined before the official discontinuation. Rather than causing a shift in the probability of survival, the gradual discontinuation would cause a change in its slope. In the RKD analysis we also found no evidence of an effect of the BCG vaccine discontinuation on the rate of change in the probability of survival in the total and female populations (Table 3). Among men, we found evidence of a slight decrease in the rate of change (−0.10 (−0.24 – 0.00)) of mortality. However, although statistically precise, the magnitude of this decrease is extremely small and thus of limited policy significance. Third, to address this same issue and the fact that more individuals born in 1975 were born after the discontinuation,(Romanus, 1983) we set the cut-off to be one year earlier (1975 in the RDD and 1974 in the multi-period DiD), which produced consistent results with those of the main analysis (Tables S 2 to S 4, Figures S 7 to S 9). Fourth, in the RDD analysis, the point estimate and inference might depend on the bandwidth within which the LATE is estimated. Increasing the bandwidth will reduce variance as more observations are included in the analysis. However, it increases the bias because these observations are less similar to each other than those closer to the cut-off. Decreasing the bandwidth would have the opposite effect and increase variance while reducing the bias. It was not possible to decrease the bandwidth due to the limited number of mass points in our analysis but increasing the bandwidth by one, two, and three years, did not change the results and all point estimates remained statistically insignificant (Table S 5).

**Table 3:** Regression Kink Design estimation results

|  | Total |  | Men |  | Women |  |
| --- | --- | --- | --- | --- | --- | --- |
|  | below<br>cut-off | above<br>cut-off | below<br>cut-off | above<br>cut-off | below<br>cut-off | above<br>cut-off |
| Sample size | 323,169 | 383,905 | 339,754 | 343,064 | 103,683 | 139,722 |
| Bandwidth | 3.56 | 3.56 | 5.01 | 5.01 | 2.83 | 2.83 |
| Local average treatment effect (percentage points) |  |  |  |  |  |  |
| Coefficient | -0.06 |  | -0.10 |  | -0.01 |  |
| 95% CI | (-0.20 - 0.07) |  | (-0.24 - 0.00) |  | (-0.31 - 0.03) |  |
| p-value | 0.35 |  | 0.05 |  | 0.78 |  |

*Note: We estimated the Regression Kink Design model by fitting a local linear trend including triangular Kernel weights, a data-driven bandwidth selection, and a regularization term. The derivation of the optimal bandwidth can result in a decimal. In line with Cattaneo et al. (2019), we used the floor of the decimal. The table displays the local average treatment effect for the birth cohort of 1976 and confidence intervals resulting from robust bias-corrected standard errors.*

## Discussion

There is continued interest in the non-specific effect of the BCG vaccine and whether populations should be vaccinated even if there is a low burden of TB (Goodridge et al., 2016). If the BCG vaccine had non-specific effects and increased adult survival among the general population, it could be a simple and cost-effective intervention to improve population health. Mechanistically, such an effect seems plausible since the BCG vaccine is thought to improve general childhood immunity (Cirovic et al., 2020). Other studies focused on other early life conditions also find evidence of life course consequences to changes in early life immunity and health (Almond, Currie, & Duque, 2018; Almond & Mazumder, 2005). Using population data and two complementary quasi-experimental methods that take advantage of the sudden discontinuation of the mandatory BCG vaccination in Sweden, we find no evidence for non-specific effects of the BCG vaccine on the probability of survival to age 30. In other words, up to this age and at the population level, the BCG vaccine does not appear to have substantial protective effects on mortality from diseases other than TB. This result is robust to multiple sensitivity and robustness checks and remains valid when estimating the effect separately for the male and female subpopulations.

Our results are in stark contrast to a previous study from Copenhagen, Denmark, which found that BCG vaccination was associated with an approximately 50% lower probability of survival (Rieckmann, Villumsen, Sørup, et al., 2017). The differences between our studies are likely due to residual confounding. In particular, the authors compare children that did and did not receive the vaccination during a period in Denmark when BCG vaccination became voluntary. Thus, parents self-selected their children into vaccination, and it is likely that the decision to vaccinate is related to characteristics that also influence the probability of survival. Such factors could include parental knowledge about the health of their child (as mentioned previously, BCG vaccination can have strong side-effects for less healthy children) and the background and socioeconomic status of parents who decide to vaccinate their children compared to those that do not. The authors undertook several steps to adjust for such potential sources of confounding; however, there is still a likelihood that the results were driven by unobserved systematic differences between the vaccinated and unvaccinated that affect both the vaccination status and survival since vaccination status was related to a direct parental choice and not a mandatory law as in our study.

The potential for non-specific effects among some sub-populations raises the question of whether vaccinating an entire population would provide a cost-effective way of realizing all potential non-specific effects without having to target specific population groups. However, such an approach would come with trade-offs. First, the intensity of the non-specific effects will depend on when the vaccine is administered. For example, Giaramellos-Bourboulis et al. (2020) suggest that the BCG vaccine, if given just before hospital discharge, increases time to first infection among the elderly. This non-specific effect may not have materialized if the vaccine had been given during childhood because adaptions in the innate immune system are only transitory. Thus, to reap all non-specific benefits, the population would have to be revaccinated in regular intervals. Second, the BCG vaccine is cost-effective against TB under standard thresholds. However, given the lack of a detectable population-level non-specific effect, it is unlikely to be cost-effective when the objective is to improve population survival in a setting like Sweden. Third, the BCG vaccine can have non-negligible side effects. While the most common side effects are mild and temporary (fever, headache, or soreness on the location where the vaccine was given), less common and more severe side effects are abscesses, bone inflammation, or disseminated TB (Raviglione Mario C., 2018). The latter is a particular risk for those who are immunocompromised. Taken together, while BCG vaccination may protect sub-populations, a broad population policy of continued vaccination is unlikely to meet conventional cost-effectiveness thresholds and comes with the risk of potentially serious side effects among individuals with a low expected benefit.

One important consideration is that our results are for Sweden, a country with a near negligible burden of TB and where infectious disease mortality, excluding deaths from Covid-19, is extremely low (The National Board of Health and Welfare, Sweden [Socialstyrelsen], 2019). The lack of a strong relationship between early childhood BCG vaccination and adult mortality in this context is not necessarily evidence that changes to early life immunity do not affect adult mortality risk, but rather that such life-course effects may become less important as the general level of infectious mortality in a population decreases. BCG vaccination may have beneficial non-specific effects for individuals at elevated risk of mortality from infectious diseases, such as those in many low- and middle-income countries or for specific at-risk groups within low-TB countries. Evaluating the non-specific effects of BCG vaccination among such populations is an important area of future work.

A second important consideration with RDD analyses is that the effect estimate of a treatment or policy applies only to those within the bandwidth around the threshold point. In our case, the RDD estimates the effect of BCG discontinuation on the first birth cohort to not receive the vaccine. If the BCG vaccine protected against infectious diseases, in particular during childhood as suggested by previous studies, there might be a time lag between the discontinuation of the vaccine and the realization of the non-specific effects as younger cohorts become exposed to larger shares of unvaccinated individuals (Biering-Sørensen et al., 2018; Giamarellos-Bourboulis et al., 2020). In this case, cohorts born after infectious diseases have crossed a critical prevalence threshold would be affected by the BCG discontinuation whereas those born during or just after the discontinuation would not. Another reason for lagged effects would be a gradual decrease in the BCG coverage over several birth cohorts rather than an abrupt change. However, both these scenarios would be captured in our multi-period DiD analysis. Therefore, the null results in both the RDD and the multi-period DiD suggest that there were no immediate nor lagged non-specific effects of the BCG vaccine.

Our study has several limitations and considerations. If an exogenous factor, which we did not account for in the analysis, caused a decline (increase) in the probability of survival at the same time as the discontinuation of BCG vaccination, it could have offset potential beneficial (harmful) non-specific effects of the BCG vaccine. To rule this out, we undertook several steps. First, in our sensitivity analysis, we varied the cut-off year. The conclusions of both quasi-experimental method analyses remain unaffected by this change. Second, we conducted an extensive search to identify potential large-scale changes in the health, economic, and political sectors in Sweden and Norway. We found no evidence of any changes in the living situation in both countries, which could have differentially affected survival at the population level in either of the countries. Another aspect to consider is that the vaccinia vaccine was discontinued in Sweden in 1976, i.e. shortly after the BCG vaccine (The Public Health Agency of Sweden, 2020). However, given the fact that the vaccinia vaccine was discontinued around the same time in Norway and we also find no statistically significant effect in the multi-period DiD analysis, we do not expect the vaccinia discontinuation to have biased our results (Norwegian Institute of Public Health, 2010). Because of data constraints, we only estimated the effect of the BCG discontinuation on survival to age 30. Including older ages, in particular the elderly where infectious diseases are more detrimental, might yield additional insights. For the same data limitations, our analysis does not assess whether the BCG vaccine protects against COVID-19 because cohort death rates are only available up to 2018.

Our study finds that the BCG vaccination did not have non-specific effects on long-term survival in Sweden. Thus, our research suggests that re-introducing or continuing national mandatory BCG vaccination in countries with similar population mortality and morbidity profiles as Sweden is unlikely to lead to substantial or cost-effective improvements in population health.

## Data Availability

All data produced will be made available online at a data repository after the publication of the article in a peer- reviewed journal.

## Supplementary Information for

Does early childhood BCG vaccination improve survival into adulthood in a population with a low tuberculosis prevalence? Quasi-experimental evidence on non-specific effects from 39 Swedish birth cohorts

**Figure S 1:**
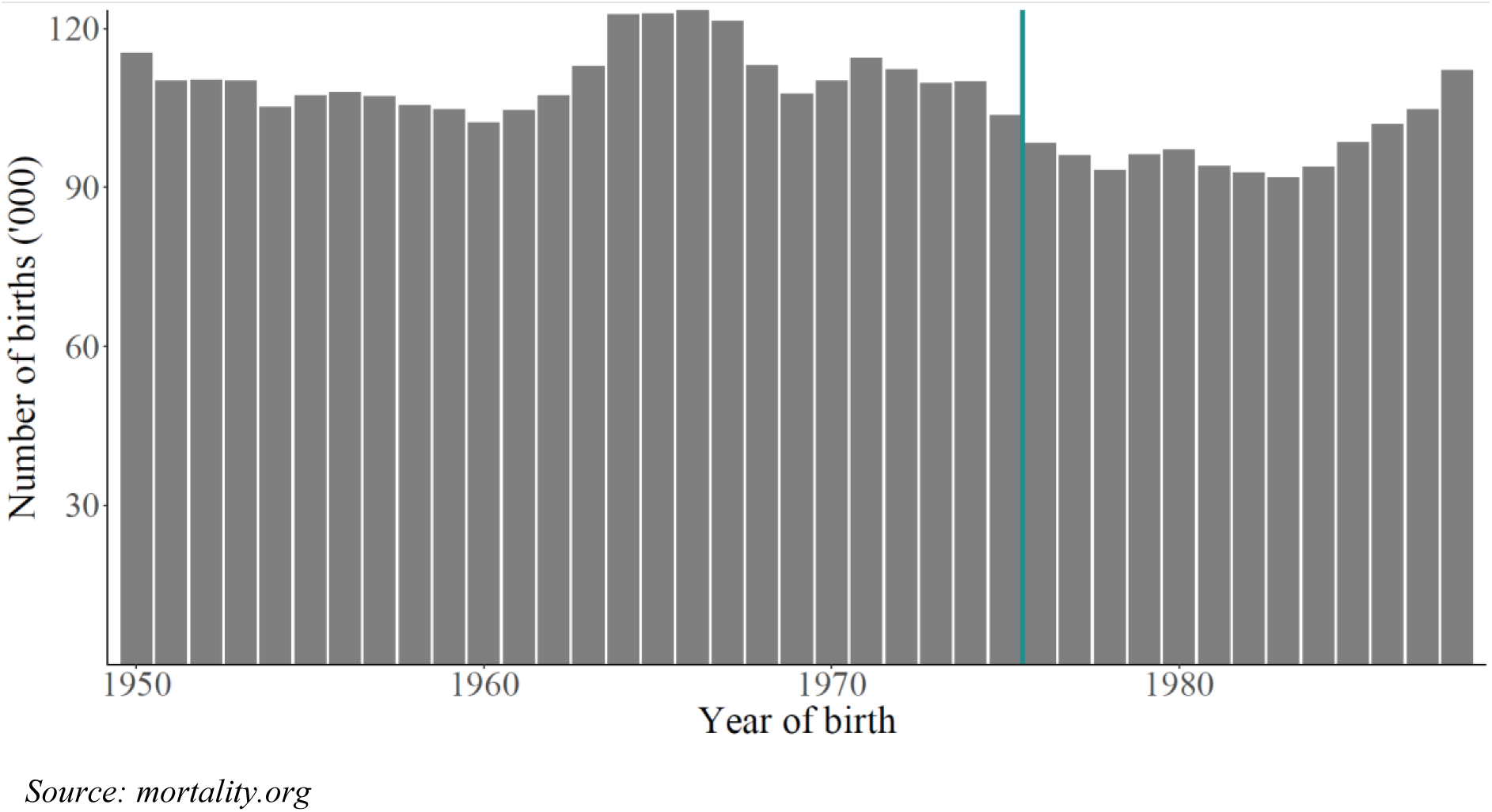
Number of births (in thousands), years 1950 - 1988

**Figure S 2:**
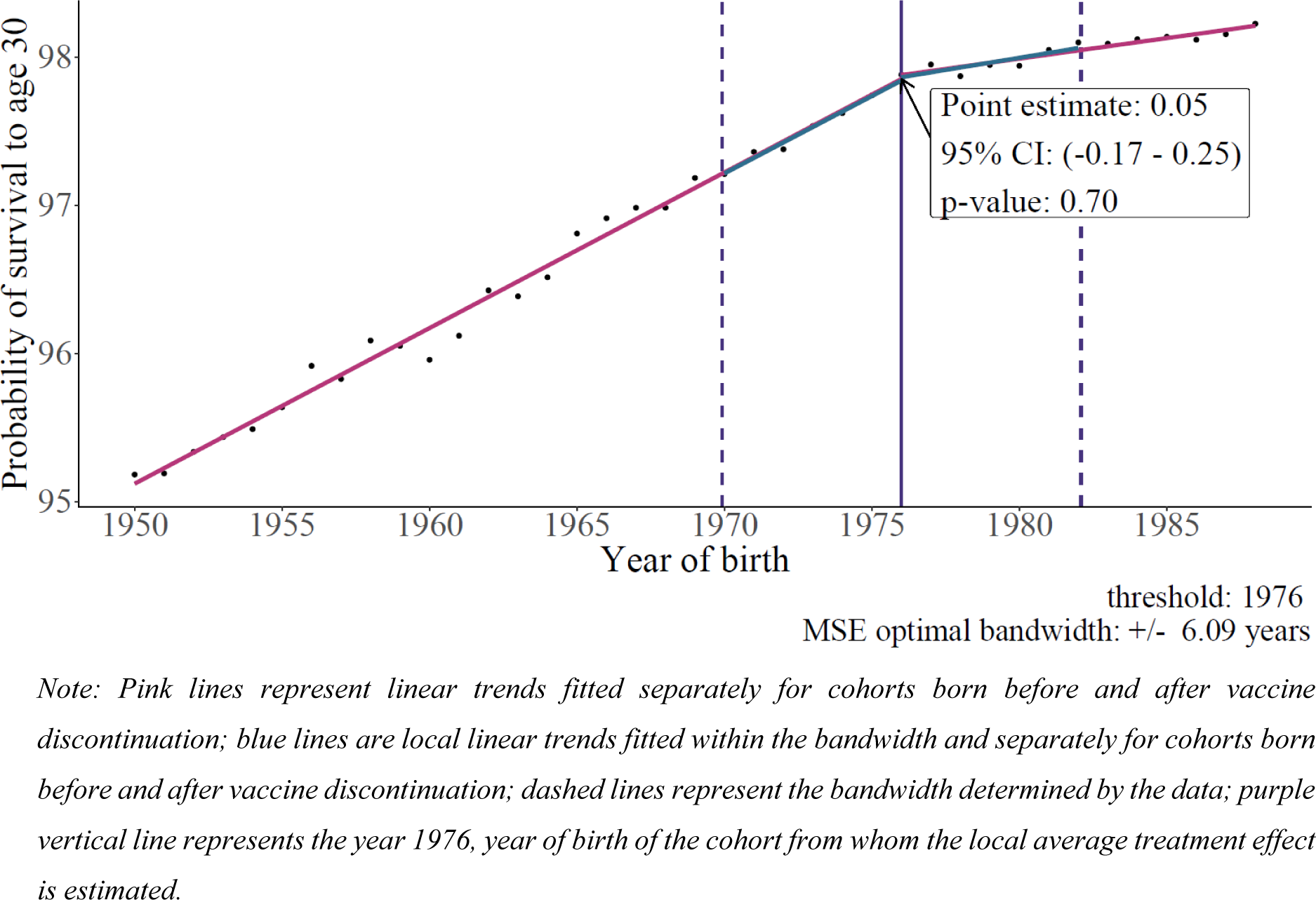
RDD: Non-specific effects of the BCG vaccine on long-term survival, male population: Note: Pink lines represent linear trends fitted separately for cohorts born before and after vaccine discontinuation; blue lines are local linear trends fitted within the bandwidth and separately for cohorts born before and after vaccine discontinuation; dashed lines represent the bandwidth determined by the data; purple vertical line represents the year 1976, year of birth of the cohort from whom the local average treatment effect is estimated.

**Figure S 3:**
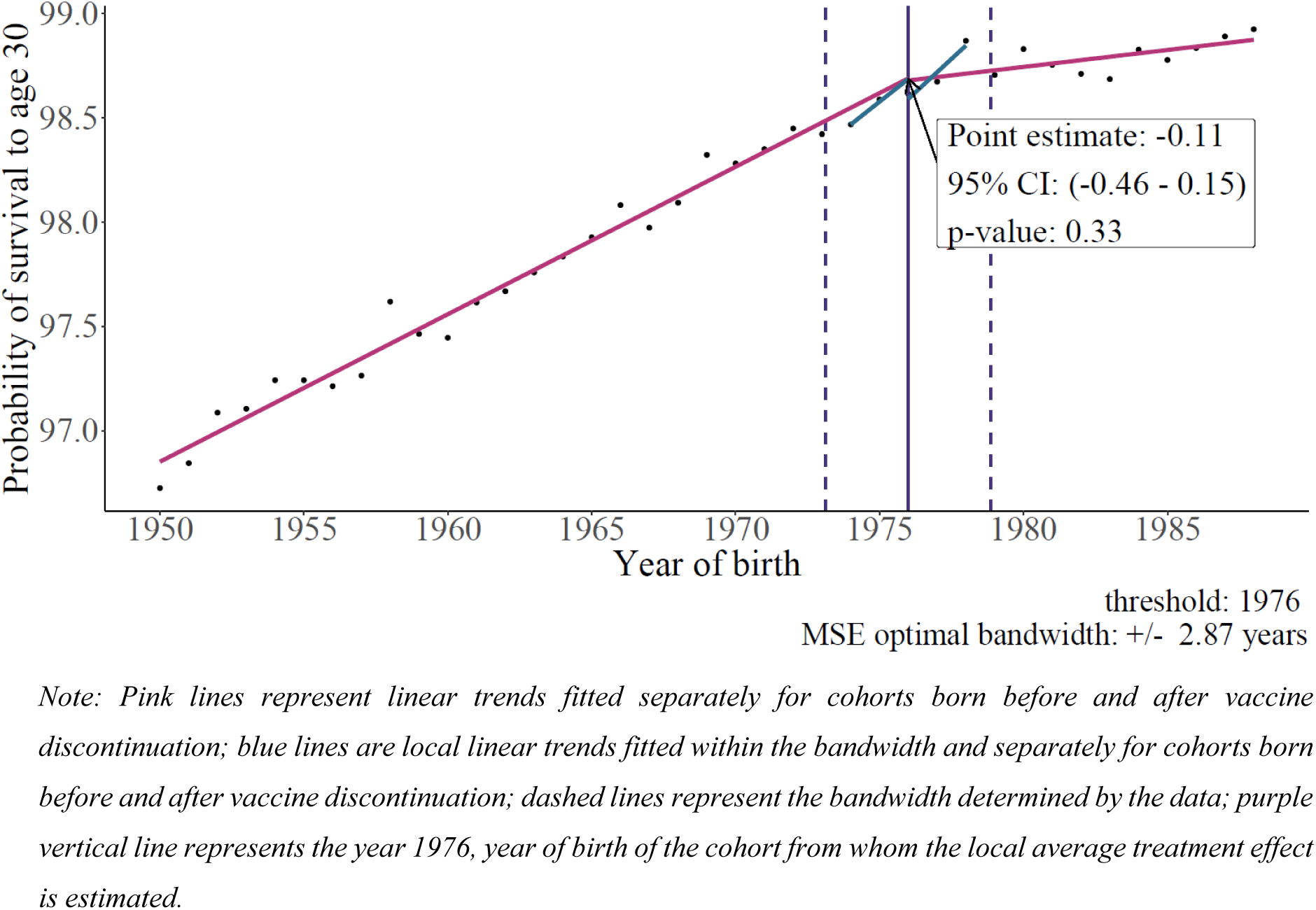
RDD: Non-specific effects of the BCG vaccine on long-term survival, female population. Note: Pink lines represent linear trends fitted separately for cohorts born before and after vaccine discontinuation; blue lines are local linear trends fitted within the bandwidth and separately for cohorts born before and after vaccine discontinuation; dashed lines represent the bandwidth determined by the data; purple vertical line represents the year 1976, year of birth of the cohort from whom the local average treatment effect is estimated.

**Figure S 4:**
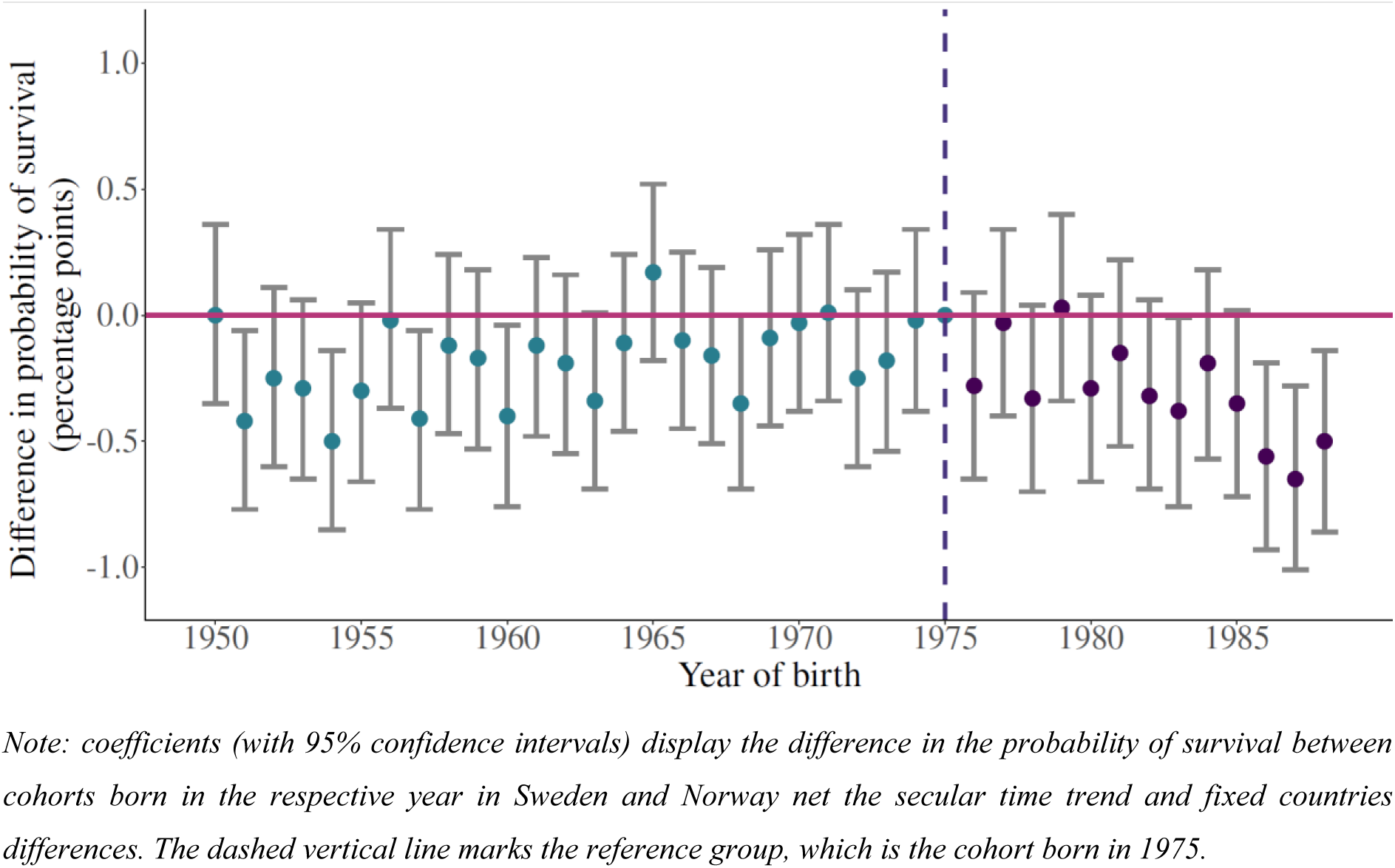
Event study plot displaying multi-period difference in differences results, male population. Note: coefficients (with 95% confidence intervals) display the difference in the probability of survival between cohorts born in the respective year in Sweden and Norway net the secular time trend and fixed countries differences. The dashed vertical line marks the reference group, which is the cohort born in 1975.

**Figure S 5:**
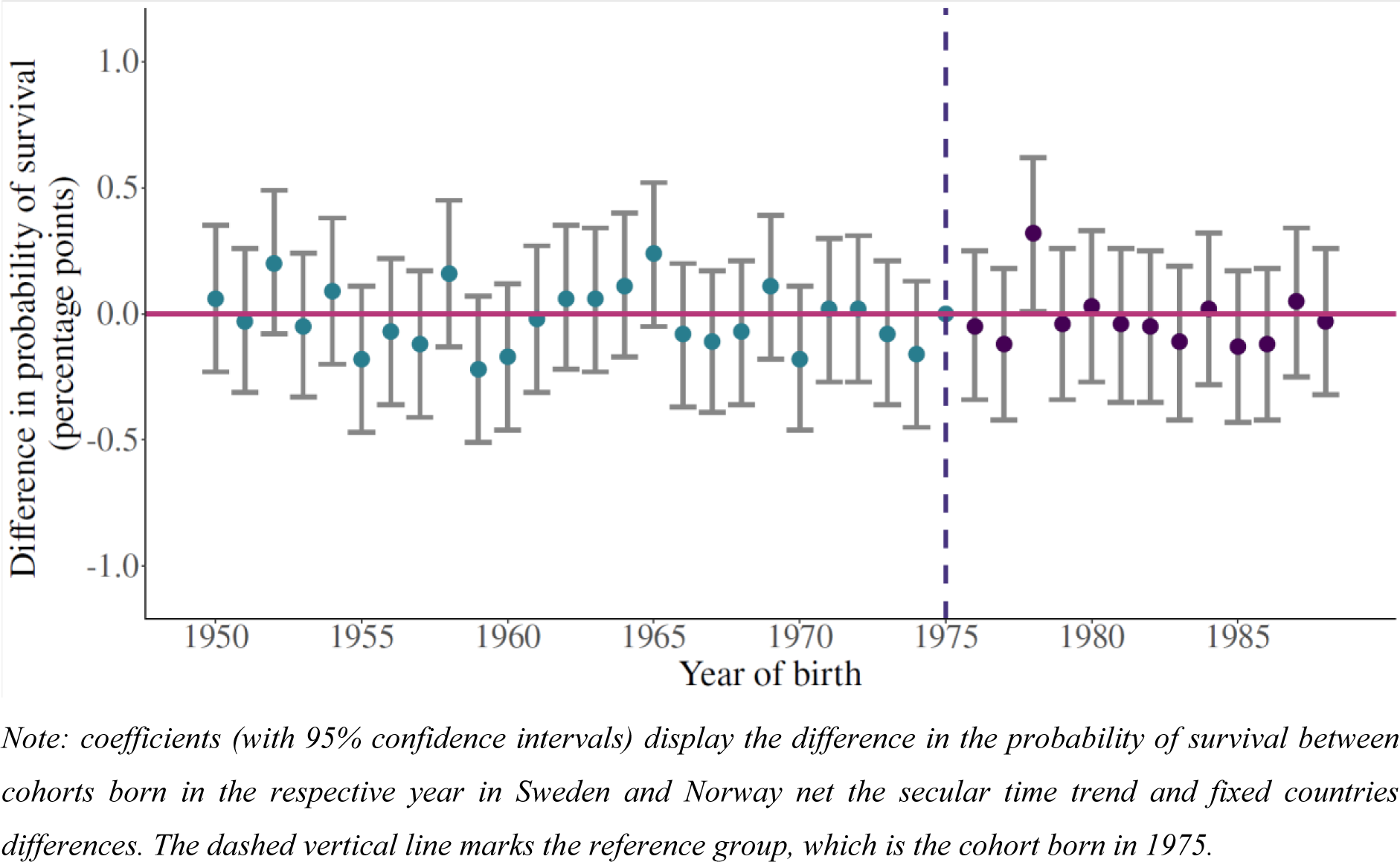
Event study plot displaying multi-period difference in differences results, female population. Note: coefficients (with 95% confidence intervals) display the difference in the probability of survival between cohorts born in the respective year in Sweden and Norway net the secular time trend and fixed countries differences. The dashed vertical line marks the reference group, which is the cohort born in 1975.

**Figure S 6:**
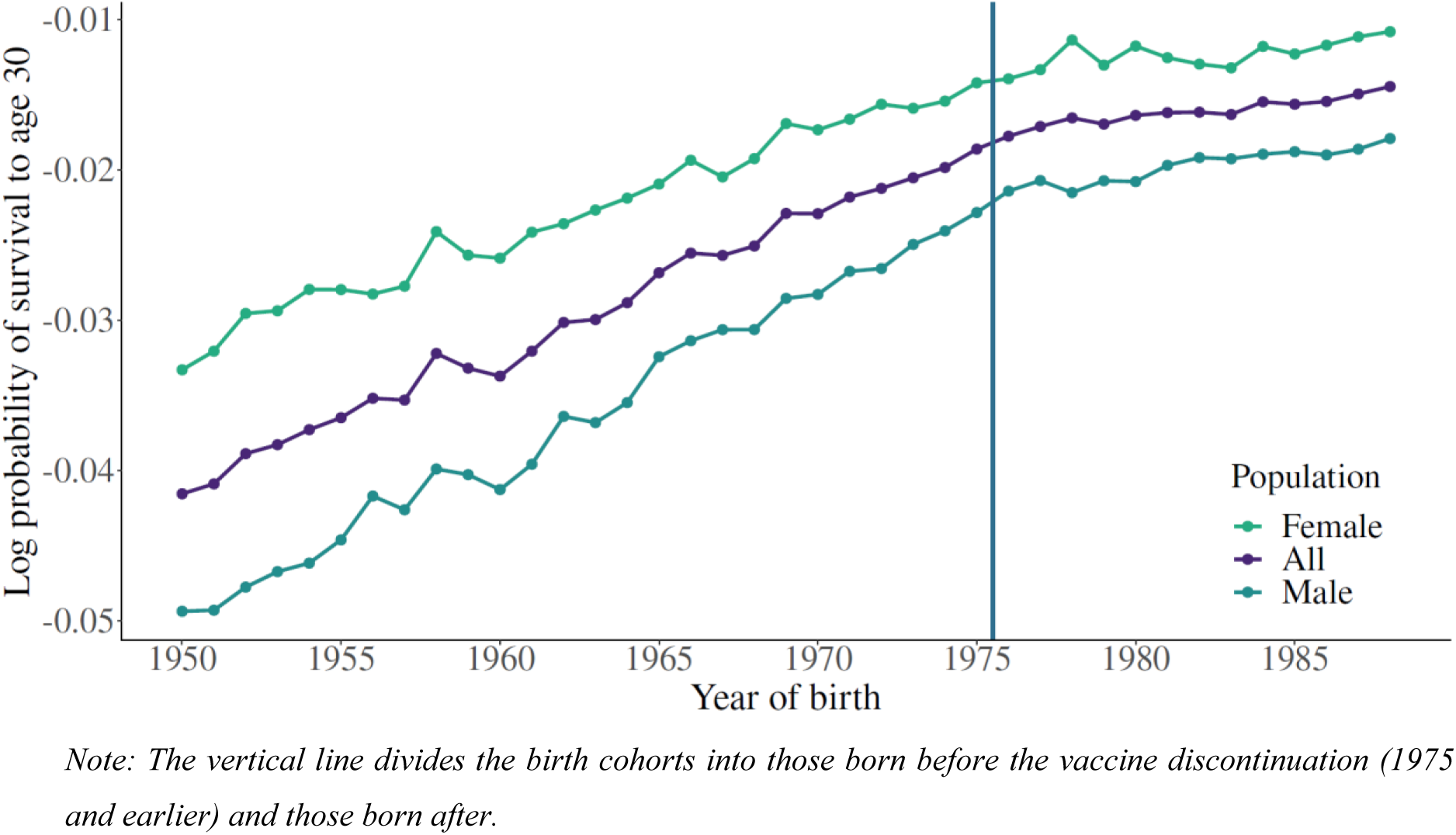
Log probability of survival to age 30 in Sweden. Note: The vertical line divides the birth cohorts into those born before the vaccine discontinuation (1975 and earlier) and those born after.

**Figure S 7:**
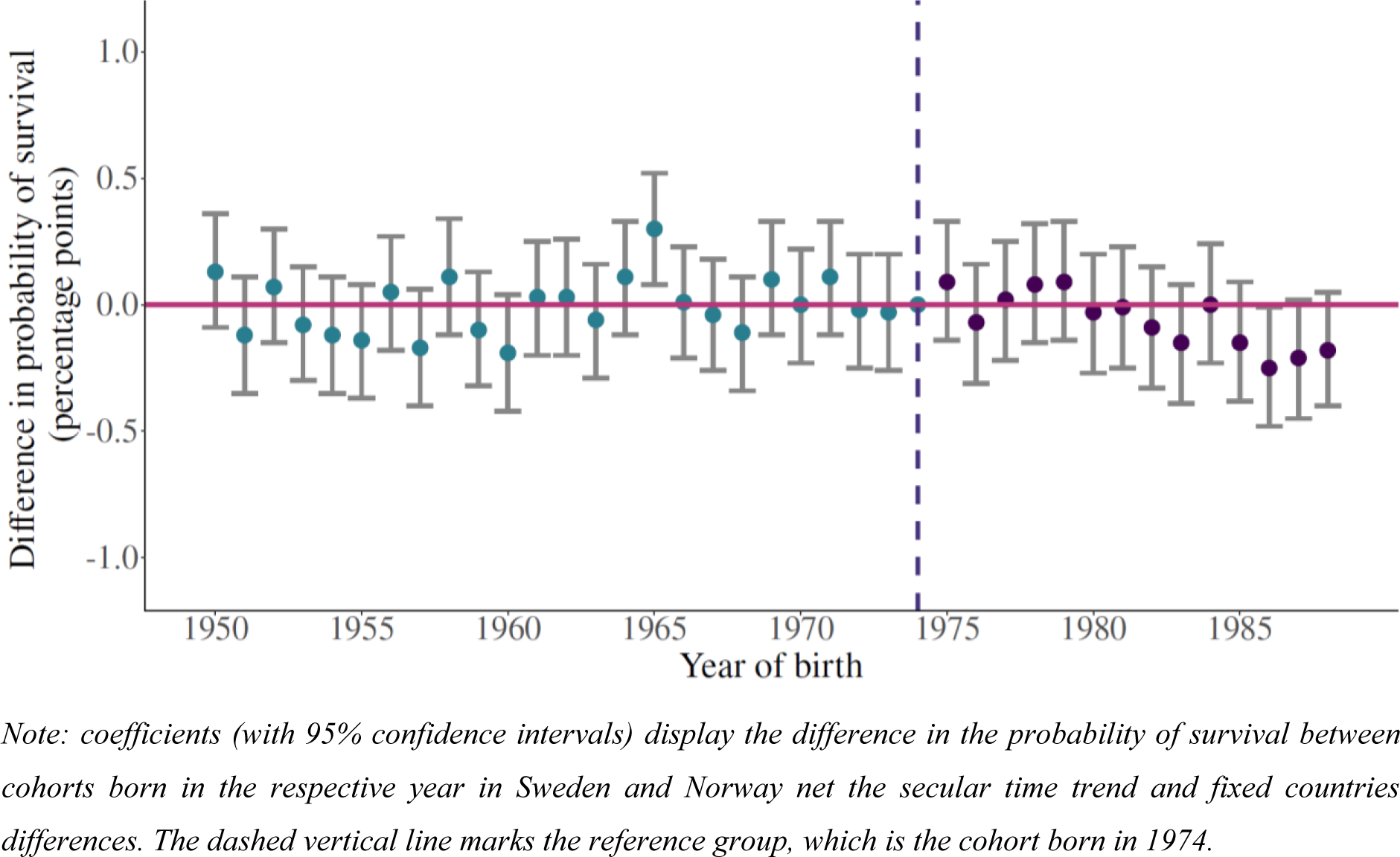
Event study plot displaying multi-period difference in differences results; reference cohort: 1974. Note: coefficients (with 95% confidence intervals) display the difference in the probability of survival between cohorts born in the respective year in Sweden and Norway net the secular time trend and fixed countries differences. The dashed vertical line marks the reference group, which is the cohort born in 1974.

**Figure S 8:**
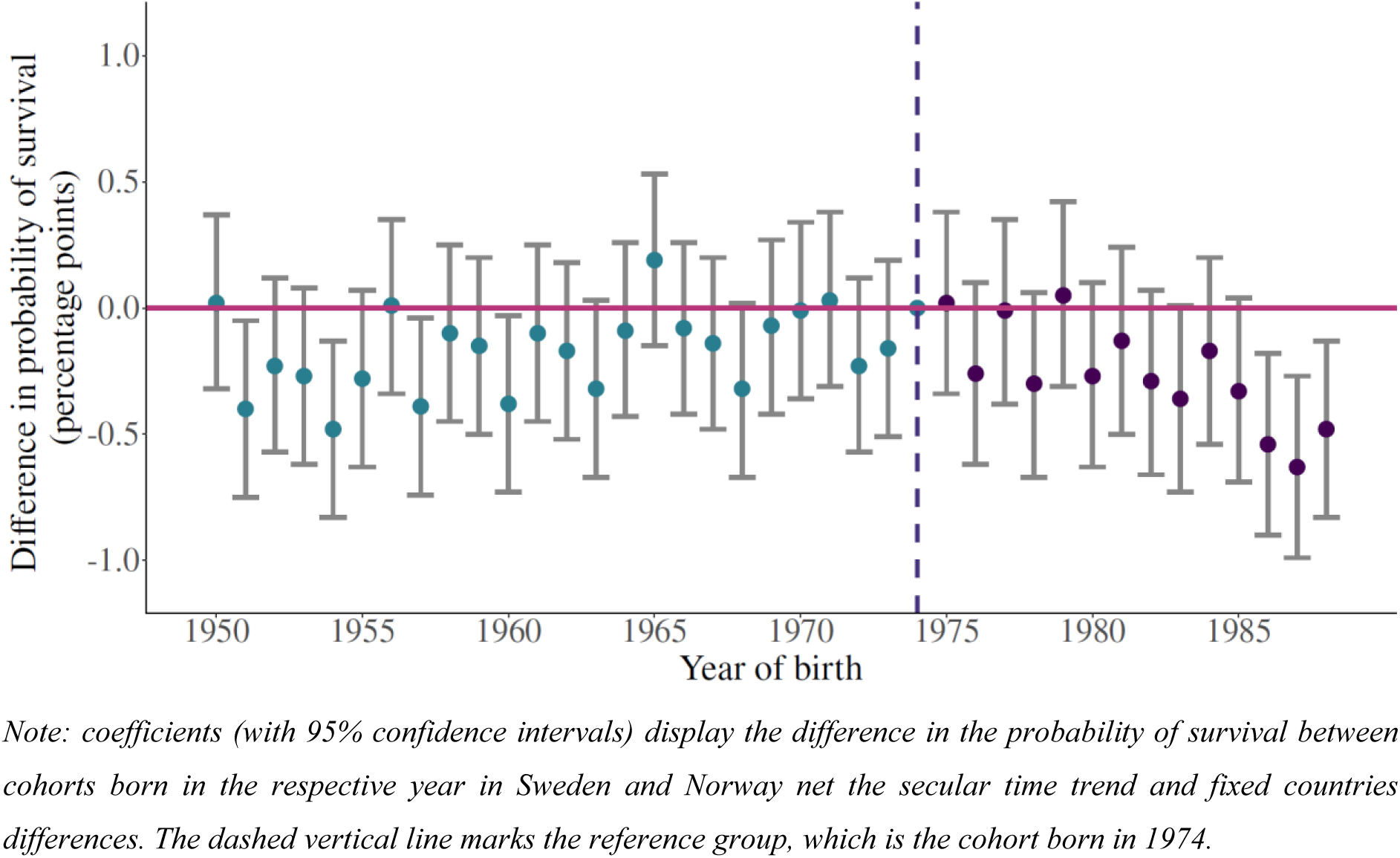
Event study plot displaying multi-period difference in differences results, male population; reference cohort: 1974. *Note: coefficients (with 95% confidence intervals) display the difference in the probability of survival between cohorts born in the respective year in Sweden and Norway net the secular time trend and fixed countries differences. The dashed vertical line marks the reference group, which is the cohort born in 1974*.

**Figure S 9:**
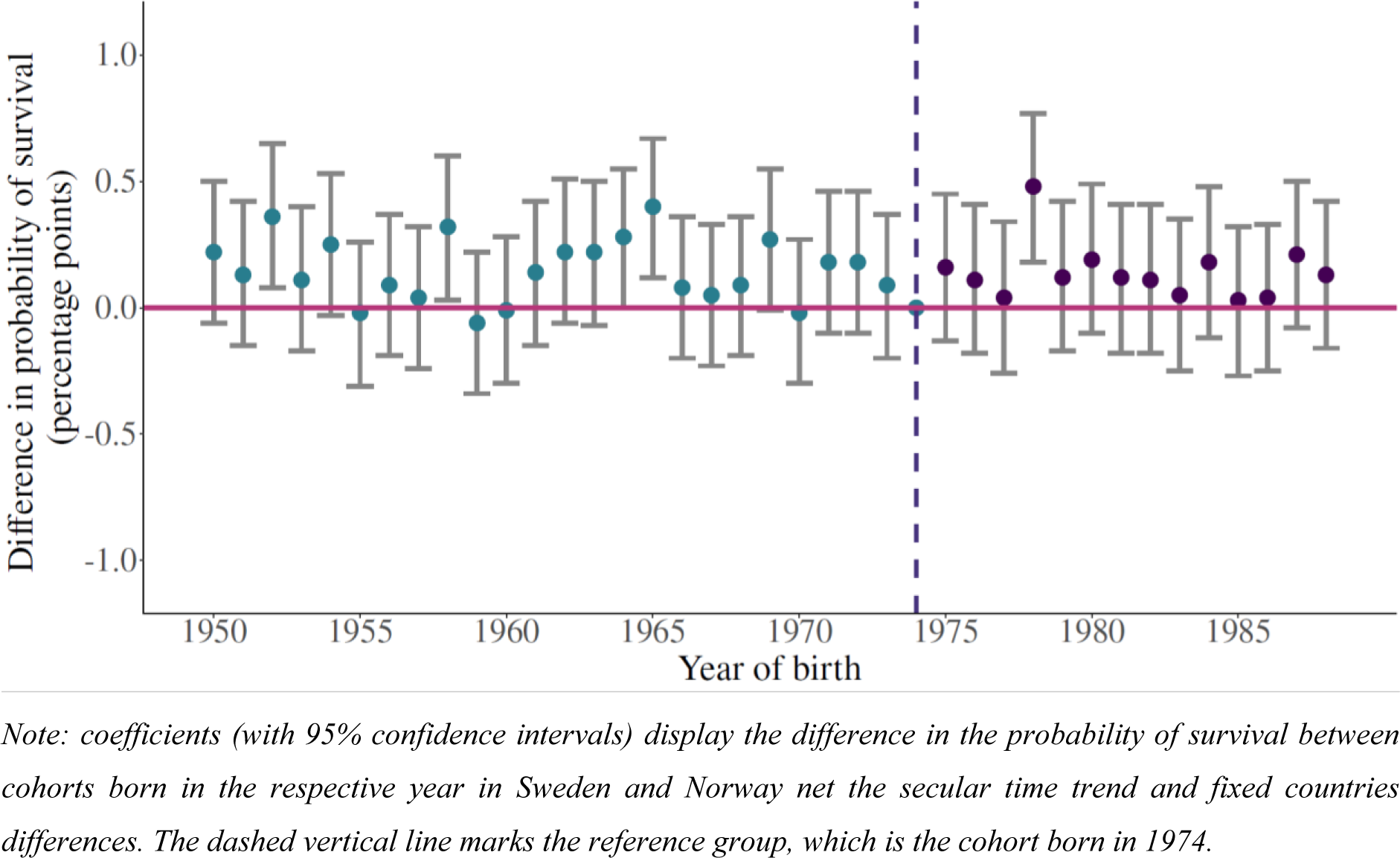
Event study plot displaying multi-period difference in differences results, female population; reference cohort: 1974. *Note: coefficients (with 95% confidence intervals) display the difference in the probability of survival between cohorts born in the respective year in Sweden and Norway net the secular time trend and fixed countries differences. The dashed vertical line marks the reference group, which is the cohort born in 1974*.

**Table S 1:**
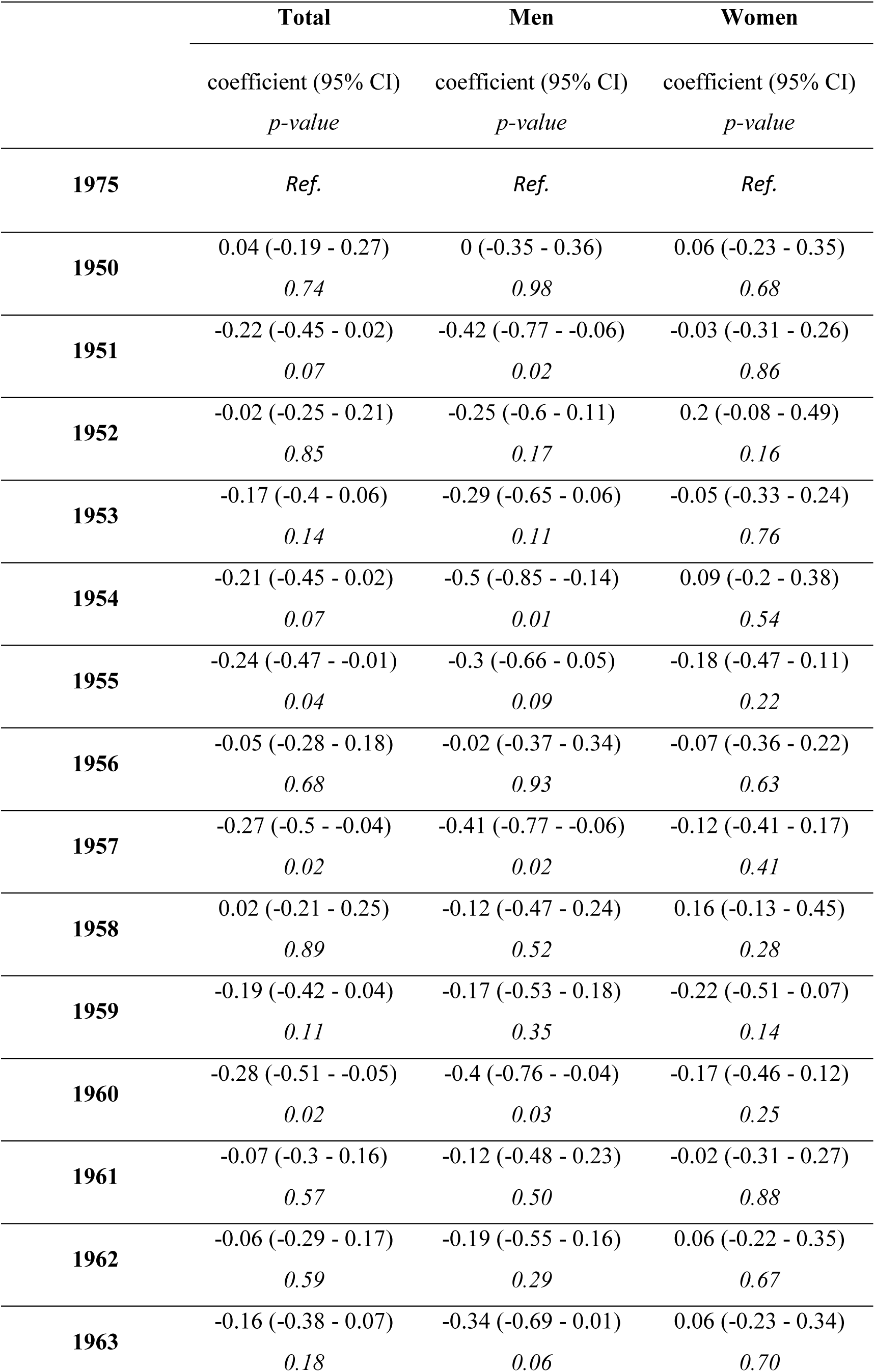
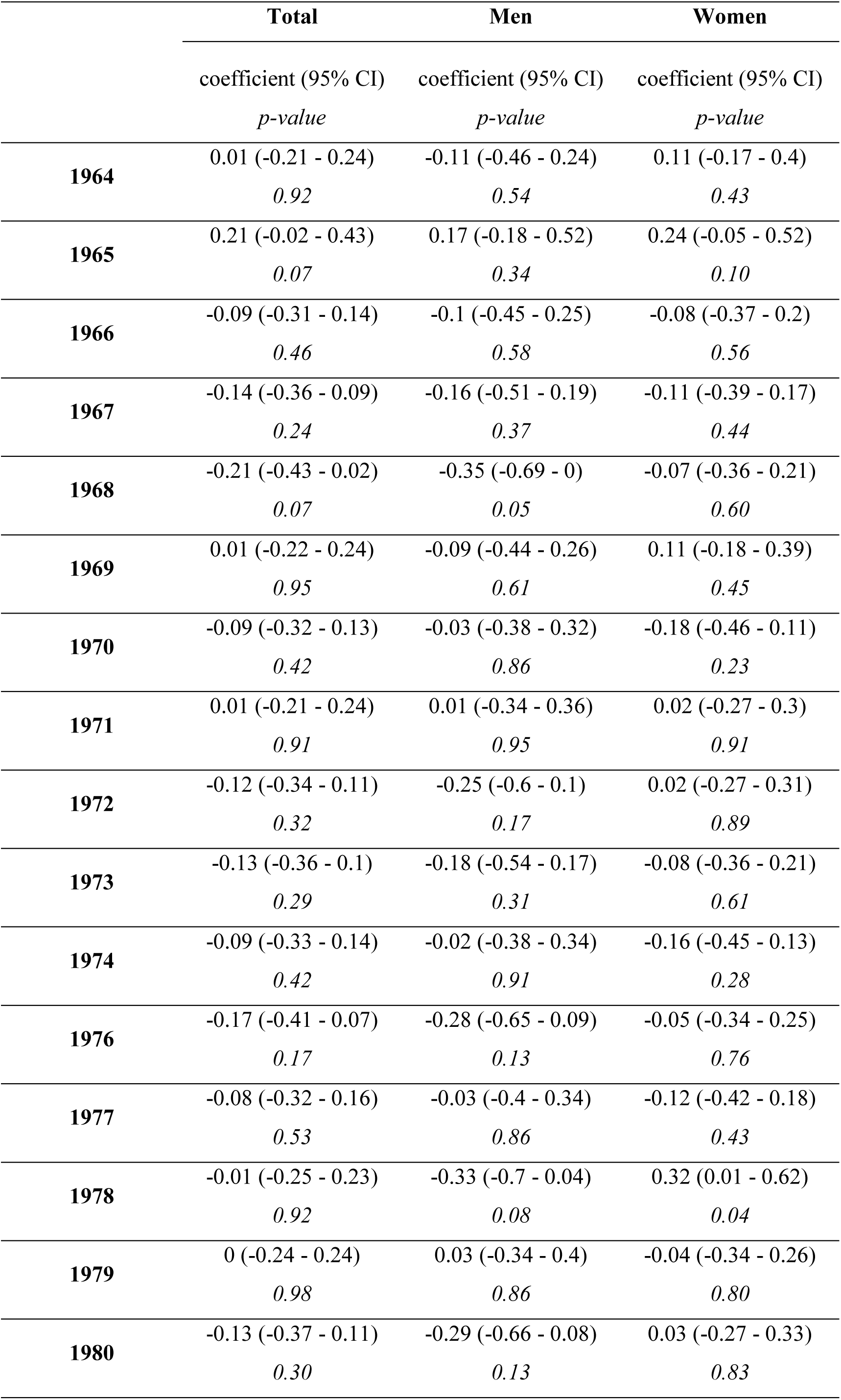
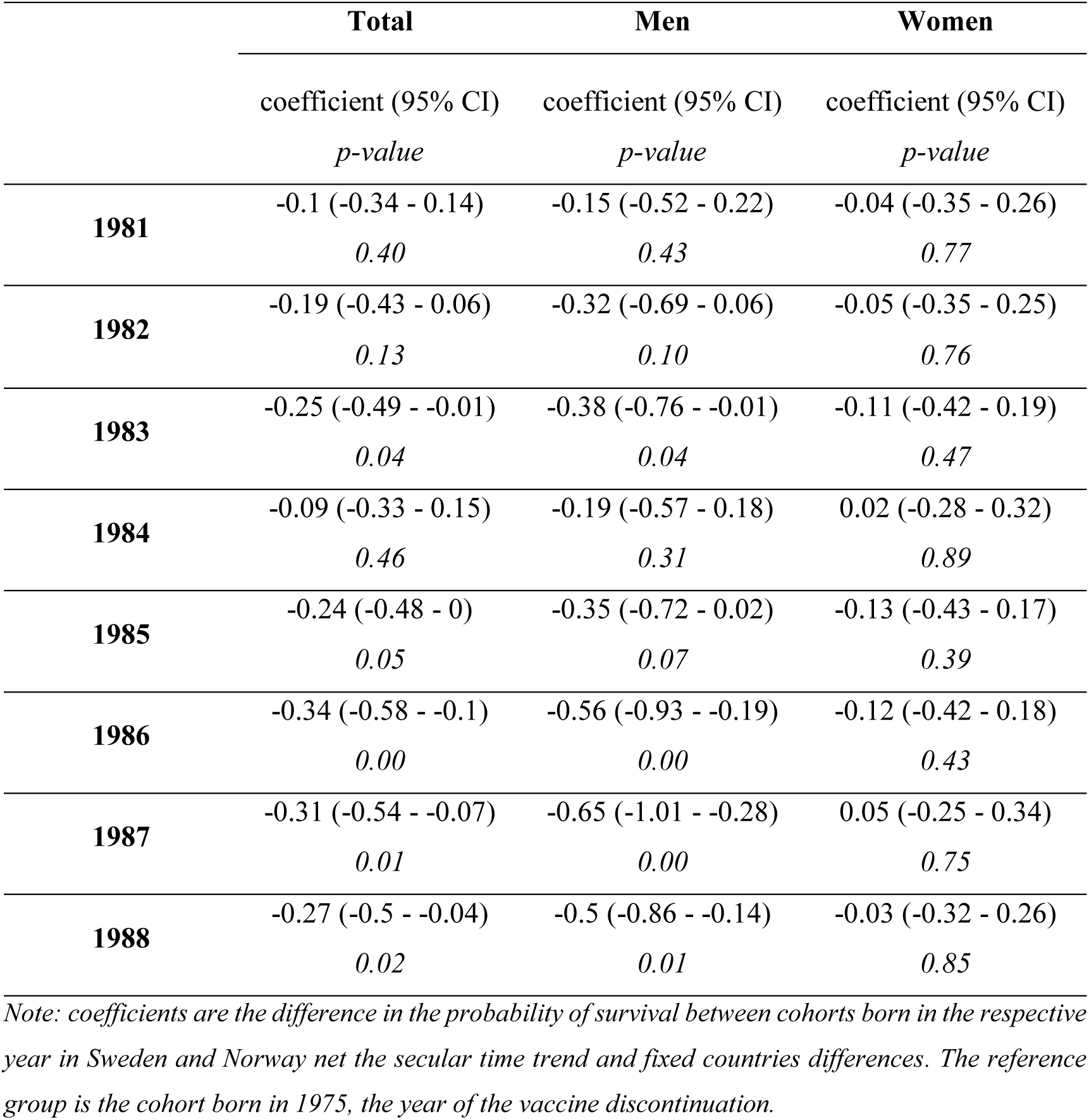
Multi-period difference in differences estimation results

|  | **Total** | **Men** | **Women** |
| --- | --- | --- | --- |
|  | coefficient (95% CI)<br>*p-value* | coefficient (95% CI)<br>*p-value* | coefficient (95% CI)<br>*p-value* |
| **1975** | *Ref.* | *Ref.* | *Ref.* |
| **1950** | 0.04 (-0.19 - 0.27)<br>0.74 | 0 (-0.35 - 0.36)<br>0.98 | 0.06 (-0.23 - 0.35)<br>0.68 |
| **1951** | -0.22 (-0.45 - 0.02)<br>0.07 | -0.42 (-0.77 - -0.06)<br>0.02 | -0.03 (-0.31 - 0.26)<br>0.86 |
| **1952** | -0.02 (-0.25 - 0.21)<br>0.85 | -0.25 (-0.6 - 0.11)<br>0.17 | 0.2 (-0.08 - 0.49)<br>0.16 |
| **1953** | -0.17 (-0.4 - 0.06)<br>0.14 | -0.29 (-0.65 - 0.06)<br>0.11 | -0.05 (-0.33 - 0.24)<br>0.76 |
| **1954** | -0.21 (-0.45 - 0.02)<br>0.07 | -0.5 (-0.85 - -0.14)<br>0.01 | 0.09 (-0.2 - 0.38)<br>0.54 |
| **1955** | -0.24 (-0.47 - -0.01)<br>0.04 | -0.3 (-0.66 - 0.05)<br>0.09 | -0.18 (-0.47 - 0.11)<br>0.22 |
| **1956** | -0.05 (-0.28 - 0.18)<br>0.68 | -0.02 (-0.37 - 0.34)<br>0.93 | -0.07 (-0.36 - 0.22)<br>0.63 |
| **1957** | -0.27 (-0.5 - -0.04)<br>0.02 | -0.41 (-0.77 - -0.06)<br>0.02 | -0.12 (-0.41 - 0.17)<br>0.41 |
| **1958** | 0.02 (-0.21 - 0.25)<br>0.89 | -0.12 (-0.47 - 0.24)<br>0.52 | 0.16 (-0.13 - 0.45)<br>0.28 |
| **1959** | -0.19 (-0.42 - 0.04)<br>0.11 | -0.17 (-0.53 - 0.18)<br>0.35 | -0.22 (-0.51 - 0.07)<br>0.14 |
| **1960** | -0.28 (-0.51 - -0.05)<br>0.02 | -0.4 (-0.76 - -0.04)<br>0.03 | -0.17 (-0.46 - 0.12)<br>0.25 |
| **1961** | -0.07 (-0.3 - 0.16)<br>0.57 | -0.12 (-0.48 - 0.23)<br>0.50 | -0.02 (-0.31 - 0.27)<br>0.88 |
| **1962** | -0.06 (-0.29 - 0.17)<br>0.59 | -0.19 (-0.55 - 0.16)<br>0.29 | 0.06 (-0.22 - 0.35)<br>0.67 |
| **1963** | -0.16 (-0.38 - 0.07)<br>0.18 | -0.34 (-0.69 - 0.01)<br>0.06 | 0.06 (-0.23 - 0.34)<br>0.70 |
|  | coefficient (95% CI) | coefficient (95% CI) | coefficient (95% CI) |
|  | *p-value* | *p-value* | *p-value* |
| **1964** | 0.01 (-0.21 - 0.24)<br>*0.92* | -0.11 (-0.46 - 0.24)<br>*0.54* | 0.11 (-0.17 - 0.4)<br>*0.43* |
| **1965** | 0.21 (-0.02 - 0.43)<br>*0.07* | 0.17 (-0.18 - 0.52)<br>*0.34* | 0.24 (-0.05 - 0.52)<br>*0.10* |
| **1966** | -0.09 (-0.31 - 0.14)<br>*0.46* | -0.1 (-0.45 - 0.25)<br>*0.58* | -0.08 (-0.37 - 0.2)<br>*0.56* |
| **1967** | -0.14 (-0.36 - 0.09)<br>*0.24* | -0.16 (-0.51 - 0.19)<br>*0.37* | -0.11 (-0.39 - 0.17)<br>*0.44* |
| **1968** | -0.21 (-0.43 - 0.02)<br>*0.07* | -0.35 (-0.69 - 0)<br>*0.05* | -0.07 (-0.36 - 0.21)<br>*0.60* |
| **1969** | 0.01 (-0.22 - 0.24)<br>*0.95* | -0.09 (-0.44 - 0.26)<br>*0.61* | 0.11 (-0.18 - 0.39)<br>*0.45* |
| **1970** | -0.09 (-0.32 - 0.13)<br>*0.42* | -0.03 (-0.38 - 0.32)<br>*0.86* | -0.18 (-0.46 - 0.11)<br>*0.23* |
| **1971** | 0.01 (-0.21 - 0.24)<br>*0.91* | 0.01 (-0.34 - 0.36)<br>*0.95* | 0.02 (-0.27 - 0.3)<br>*0.91* |
| **1972** | -0.12 (-0.34 - 0.11)<br>*0.32* | -0.25 (-0.6 - 0.1)<br>*0.17* | 0.02 (-0.27 - 0.31)<br>*0.89* |
| **1973** | -0.13 (-0.36 - 0.1)<br>*0.29* | -0.18 (-0.54 - 0.17)<br>*0.31* | -0.08 (-0.36 - 0.21)<br>*0.61* |
| **1974** | -0.09 (-0.33 - 0.14)<br>*0.42* | -0.02 (-0.38 - 0.34)<br>*0.91* | -0.16 (-0.45 - 0.13)<br>*0.28* |
| **1976** | -0.17 (-0.41 - 0.07)<br>*0.17* | -0.28 (-0.65 - 0.09)<br>*0.13* | -0.05 (-0.34 - 0.25)<br>*0.76* |
| **1977** | -0.08 (-0.32 - 0.16)<br>*0.53* | -0.03 (-0.4 - 0.34)<br>*0.86* | -0.12 (-0.42 - 0.18)<br>*0.43* |
| **1978** | -0.01 (-0.25 - 0.23)<br>*0.92* | -0.33 (-0.7 - 0.04)<br>*0.08* | 0.32 (0.01 - 0.62)<br>*0.04* |
| **1979** | 0 (-0.24 - 0.24)<br>*0.98* | 0.03 (-0.34 - 0.4)<br>*0.86* | -0.04 (-0.34 - 0.26)<br>*0.80* |
| **1980** | -0.13 (-0.37 - 0.11)<br>*0.30* | -0.29 (-0.66 - 0.08)<br>*0.13* | 0.03 (-0.27 - 0.33)<br>*0.83* |
|  | coefficient (95% CI) | coefficient (95% CI) | coefficient (95% CI) |
|  | *p-value* | *p-value* | *p-value* |
| **1981** | -0.1 (-0.34 - 0.14)<br>*0.40* | -0.15 (-0.52 - 0.22)<br>*0.43* | -0.04 (-0.35 - 0.26)<br>*0.77* |
| **1982** | -0.19 (-0.43 - 0.06)<br>*0.13* | -0.32 (-0.69 - 0.06)<br>*0.10* | -0.05 (-0.35 - 0.25)<br>*0.76* |
| **1983** | -0.25 (-0.49 - -0.01)<br>*0.04* | -0.38 (-0.76 - -0.01)<br>*0.04* | -0.11 (-0.42 - 0.19)<br>*0.47* |
| **1984** | -0.09 (-0.33 - 0.15)<br>*0.46* | -0.19 (-0.57 - 0.18)<br>*0.31* | 0.02 (-0.28 - 0.32)<br>*0.89* |
| **1985** | -0.24 (-0.48 - 0)<br>*0.05* | -0.35 (-0.72 - 0.02)<br>*0.07* | -0.13 (-0.43 - 0.17)<br>*0.39* |
| **1986** | -0.34 (-0.58 - -0.1)<br>*0.00* | -0.56 (-0.93 - -0.19)<br>*0.00* | -0.12 (-0.42 - 0.18)<br>*0.43* |
| **1987** | -0.31 (-0.54 - -0.07)<br>*0.01* | -0.65 (-1.01 - -0.28)<br>*0.00* | 0.05 (-0.25 - 0.34)<br>*0.75* |
| **1988** | -0.27 (-0.5 - -0.04)<br>*0.02* | -0.5 (-0.86 - -0.14)<br>*0.01* | -0.03 (-0.32 - 0.26)<br>*0.85* |

*Note: coefficients are the difference in the probability of survival between cohorts born in the respective year in Sweden and Norway net the secular time trend and fixed countries differences. The reference group is the cohort born in 1975, the year of the vaccine discontinuation.*

**Table S 2:** Regression Discontinuity Design estimation results, threshold year 1975

|  | Total |  | Men |  | Women |  |
| --- | --- | --- | --- | --- | --- | --- |
|  | below<br>cut-off | above<br>cut-off | below<br>cut-off | above<br>cut-off | below<br>cut-off | above<br>cut-off |
| Sample size | 446,294 | 487,537 | 170,896 | 201,170 | 216,324 | 236,954 |
| Bandwidth | 4.03 | 4.03 | 3.37 | 3.37 | 4.79 | 4.79 |
| Local average treatment effect (percentage points) |  |  |  |  |  |  |
| Coefficient | 0.06 |  | 0.02 |  | 0.09 |  |
| 95% CI | (-0.13 - 0.23) |  | (-0.31 - 0.33) |  | (-0.1 - 0.3) |  |
| p-value | 0.59 |  | 0.97 |  | 0.33 |  |

*Note: We estimated the Regression Discontinuity Design model by fitting a local linear trend including triangular Kernel weights, a data-driven bandwidth selection, and a regularization term. The derivation of the optimal bandwidth can result in a decimal. In line with Cattaneo et al. (2019), we used the floor of the decimal. The table displays the local average treatment effect for the birth cohort of 1975 and confidence intervals resulting from robust bias-corrected standard errors.*

**Table S 3:** Multi-period difference in differences estimation results, cut-off 1974

|  | **Total** | **Men** | **Women** |
| --- | --- | --- | --- |
|  | coefficient (95% CI) | coefficient (95% CI) | coefficient (95% CI) |
|  | *p-value* | *p-value* | *p-value* |
| **1974** | *Ref.* | *Ref.* | *Ref.* |
| **1950** | 0.13 (-0.09 - 0.36)<br>*0.25* | 0.02 (-0.32 - 0.37)<br>*0.89* | 0.22 (-0.06 - 0.50)<br>*0.13* |
| **1951** | -0.12 (-0.35 - 0.11)<br>*0.30* | -0.4 (-0.75 - -0.05)<br>*0.03* | 0.13 (-0.15 - 0.42)<br>*0.35* |
| **1952** | 0.07 (-0.15 - 0.3)<br>*0.53* | -0.23 (-0.57 - 0.12)<br>*0.21* | 0.36 (0.08 - 0.65)<br>*0.01* |
| **1953** | -0.08 (-0.3 - 0.15)<br>*0.50* | -0.27 (-0.62 - 0.08)<br>*0.13* | 0.11 (-0.17 - 0.4)<br>*0.43* |
| **1954** | -0.12 (-0.35 - 0.11)<br>*0.30* | -0.48 (-0.83 - -0.13)<br>*0.01* | 0.25 (-0.03 - 0.53)<br>*0.09* |
| **1955** | -0.14 (-0.37 - 0.08)<br>*0.21* | -0.28 (-0.63 - 0.07)<br>*0.11* | -0.02 (-0.31 - 0.26)<br>*0.88* |
| **1956** | 0.05 (-0.18 - 0.27)<br>*0.69* | 0.01 (-0.34 - 0.35)<br>*0.98* | 0.09 (-0.19 - 0.37)<br>*0.53* |
| **1957** | -0.17 (-0.4 - 0.06)<br>*0.14* | -0.39 (-0.74 - -0.04)<br>*0.03* | 0.04 (-0.24 - 0.32)<br>*0.79* |
| **1958** | 0.11 (-0.12 - 0.34)<br>*0.34* | -0.1 (-0.45 - 0.25)<br>*0.59* | 0.32 (0.03 - 0.6)<br>*0.03* |
| **1959** | -0.1 (-0.32 - 0.13)<br>*0.41* | -0.15 (-0.5 - 0.2)<br>*0.40* | -0.06 (-0.34 - 0.22)<br>*0.68* |
| **1960** | -0.19 (-0.42 - 0.04)<br>*0.10* | -0.38 (-0.73 - -0.03)<br>*0.04* | -0.01 (-0.3 - 0.28)<br>*0.94* |
| **1961** | 0.03 (-0.2 - 0.25)<br>*0.81* | -0.1 (-0.45 - 0.25)<br>*0.57* | 0.14 (-0.15 - 0.42)<br>*0.34* |
| **1962** | 0.03 (-0.2 - 0.26)<br>*0.79* | -0.17 (-0.52 - 0.18)<br>*0.34* | 0.22 (-0.06 - 0.51)<br>*0.12* |
| **1963** | -0.06 (-0.29 - 0.16)<br>*0.60* | -0.32 (-0.67 - 0.03)<br>*0.07* | 0.22 (-0.07 - 0.5)<br>*0.13* |
|  | coefficient (95% CI) | coefficient (95% CI) | coefficient (95% CI) |
|  | *p-value* | *p-value* | *p-value* |
| **1964** | 0.11 (-0.12 - 0.33)<br>*0.35* | -0.09 (-0.43 - 0.26)<br>*0.62* | 0.28 (0 - 0.55)<br>*0.05* |
| **1965** | 0.3 (0.08 - 0.52)<br>*0.01* | 0.19 (-0.15 - 0.53)<br>*0.28* | 0.4 (0.12 - 0.67)<br>*0.01* |
| **1966** | 0.01 (-0.21 - 0.23)<br>*0.94* | -0.08 (-0.42 - 0.26)<br>*0.65* | 0.08 (-0.2 - 0.36)<br>*0.59* |
| **1967** | -0.04 (-0.26 - 0.18)<br>*0.72* | -0.14 (-0.48 - 0.2)<br>*0.42* | 0.05 (-0.23 - 0.33)<br>*0.73* |
| **1968** | -0.11 (-0.34 - 0.11)<br>*0.32* | -0.32 (-0.67 - 0.02)<br>*0.06* | 0.09 (-0.19 - 0.36)<br>*0.55* |
| **1969** | 0.1 (-0.12 - 0.33)<br>*0.37* | -0.07 (-0.42 - 0.27)<br>*0.69* | 0.27 (-0.01 - 0.55)<br>*0.06* |
| **1970** | 0 (-0.23 - 0.22)<br>*1.00* | -0.01 (-0.36 - 0.34)<br>*0.95* | -0.02 (-0.3 - 0.27)<br>*0.91* |
| **1971** | 0.11 (-0.12 - 0.33)<br>*0.35* | 0.03 (-0.31 - 0.38)<br>*0.86* | 0.18 (-0.1 - 0.46)<br>*0.22* |
| **1972** | -0.02 (-0.25 - 0.2)<br>*0.85* | -0.23 (-0.57 - 0.12)<br>*0.20* | 0.18 (-0.1 - 0.46)<br>*0.21* |
| **1973** | -0.03 (-0.26 - 0.2)<br>*0.79* | -0.16 (-0.51 - 0.19)<br>*0.36* | 0.09 (-0.2 - 0.37)<br>*0.56* |
| **1975** | 0.09 (-0.14 - 0.33)<br>*0.42* | 0.02 (-0.34 - 0.38)<br>*0.91* | 0.16 (-0.13 - 0.45)<br>*0.28* |
| **1976** | -0.07 (-0.31 - 0.16)<br>*0.54* | -0.26 (-0.62 - 0.1)<br>*0.16* | 0.11 (-0.18 - 0.41)<br>*0.45* |
| **1977** | 0.02 (-0.22 - 0.25)<br>*0.89* | -0.01 (-0.38 - 0.35)<br>*0.95* | 0.04 (-0.26 - 0.34)<br>*0.79* |
| **1978** | 0.08 (-0.15 - 0.32)<br>*0.50* | -0.3 (-0.67 - 0.06)<br>*0.10* | 0.48 (0.18 - 0.77)<br>*0.00* |
| **1979** | 0.09 (-0.14 - 0.33)<br>*0.45* | 0.05 (-0.31 - 0.42)<br>*0.77* | 0.12 (-0.17 - 0.42)<br>*0.42* |
| **1980** | -0.03 (-0.27 - 0.2)<br>*0.78* | -0.27 (-0.63 - 0.1)<br>*0.15* | 0.19 (-0.1 - 0.49)<br>*0.20* |
|  | coefficient (95% CI) | coefficient (95% CI) | coefficient (95% CI) |
|  | *p-value* | *p-value* | *p-value* |
| **1981** | -0.01 (-0.25 - 0.23)<br>*0.94* | -0.13 (-0.5 - 0.24)<br>*0.49* | 0.12 (-0.18 - 0.41)<br>*0.44* |
| **1982** | -0.09 (-0.33 - 0.15)<br>*0.45* | -0.29 (-0.66 - 0.07)<br>*0.11* | 0.11 (-0.18 - 0.41)<br>*0.46* |
| **1983** | -0.15 (-0.39 - 0.08)<br>*0.21* | -0.36 (-0.73 - 0.01)<br>*0.05* | 0.05 (-0.25 - 0.35)<br>*0.75* |
| **1984** | 0 (-0.23 - 0.24)<br>*0.98* | -0.17 (-0.54 - 0.2)<br>*0.36* | 0.18 (-0.12 - 0.48)<br>*0.23* |
| **1985** | -0.15 (-0.38 - 0.09)<br>*0.23* | -0.33 (-0.69 - 0.04)<br>*0.08* | 0.03 (-0.27 - 0.32)<br>*0.85* |
| **1986** | -0.25 (-0.48 - -0.01)<br>*0.04* | -0.54 (-0.9 - -0.18)<br>*0.00* | 0.04 (-0.25 - 0.33)<br>*0.79* |
| **1987** | -0.21 (-0.45 - 0.02)<br>*0.07* | -0.63 (-0.99 - -0.27)<br>*0.00* | 0.21 (-0.08 - 0.5)<br>*0.16* |
| **1988** | -0.18 (-0.4 - 0.05)<br>*0.13* | -0.48 (-0.83 - -0.13)<br>*0.01* | 0.13 (-0.16 - 0.42)<br>*0.37* |

*Note: coefficients are the difference in the probability of survival between cohorts born in the respective year in Sweden and Norway net the secular time trend and fixed countries differences. As a robustness check, we defined the reference group to be the cohort born in 1974, the year before the vaccine discontinuation.*

**Table S 4:** Pooled Difference in Difference estimates, cut-off 1974

|  | Total | Male | Female |
| --- | --- | --- | --- |
| country*post | -0.05 | -0.08 | -0.02 |
|  | (-0.11 - 0.00) | (-0.16 - 0.01) | (-0.09 - 0.05) |
|  | *0.05* | *0.07* | *0.56* |

*Note: We pooled data from all pre-discontinuation and post-discontinuation years respectively. Regression models include year dummies. The table displays 95% confidence intervals in parenthesis and p-values in italic. The cut-off year was 1974.*

**Table S 5:** Regression Discontinuity Design estimation results with increasing bandwidth

|  | Total population (MSE: +/- 3.95 years) |  |  |  |  |  |
| --- | --- | --- | --- | --- | --- | --- |
|  | MSE optimal bandwidth |  | MSE optimal bandwidth |  | MSE optimal bandwidth |  |
|  | + 1 year |  | + 2 year |  | + 3 year |  |
|  | below | above | below | above | below | above |
|  | cut-off | cut-off | cut-off | cut-off | cut-off | cut-off |
| Sample size | 435,442 | 480,969 | 549,926 | 575,034 | 660,076 | 667,782 |
| Bandwidth | 4.95 | 4.95 | 5.95 | 5.95 | 6.95 | 6.95 |
|  | Local average treatment effect (percentage points) |  |  |  |  |  |
| Coefficient | 0.02 |  | 0.04 |  | 0.04 |  |
| 95% CI | (-0.30 - 0.19) |  | (-0.23 - 0.16) |  | (-0.18 - 0.17) |  |
| p-value | 0.66 |  | 0.73 |  | 0.96 |  |
|  | Male population (MSE: +/- 7.09 years) |  |  |  |  |  |
|  | MSE optimal bandwidth |  | MSE optimal bandwidth |  | MSE optimal bandwidth |  |
|  | + 1 year |  | + 2 year |  | + 3 year |  |
|  | below | above | below | above | below | above |
|  | cut-off | cut-off | cut-off | cut-off | cut-off | cut-off |
| Sample size | 395,382 | 390,377 | 453,503 | 438,717 | 516,031 | 489,465 |
| Bandwidth | 7.09 | 7.09 | 8.09 | 8.09 | 9.09 | 9.09 |
|  | Local average treatment effect (percentage points) |  |  |  |  |  |
| Coefficient | 0.04 |  | 0.05 |  | 0.04 |  |
| 95% CI | (-0.22 - 0.29) |  | (-0.20 - 0.26) |  | (-0.17 - 0.25) |  |
| p-value | 0.79 |  | 0.80 |  | 0.70 |  |

| **Female population (MSE: +/- 3.87 years)** |  |  |  |  |  |  |
| --- | --- | --- | --- | --- | --- | --- |
|  | MSE optimal bandwidth<br>+ 1 year |  | MSE optimal bandwidth<br>+ 2 year |  | MSE optimal bandwidth<br>+ 3 year |  |
|  | below<br>cut-off | above<br>cut-off | below<br>cut-off | above<br>cut-off | below<br>cut-off | above<br>cut-off |
| Sample size | 156935 | 186564 | 211304 | 233768 | 266714 | 279702 |
| Bandwidth | 3.87 | 3.87 | 4.87 | 4.87 | 5.87 | 5.87 |
| Local average treatment effect (percentage points) |  |  |  |  |  |  |
| Coefficient | -0.05 |  | 0.00 |  | 0.03 |  |
| 95% CI | (-0.65 - 0.28) |  | (-0.49 - 0.14) |  | (-0.34 - 0.16) |  |
| p-value | 0.44 |  | 0.27 |  | 0.47 |  |

*Note: We estimated the Regression Discontinuity Design model by fitting a local linear trend including triangular Kernel weights, a data-driven bandwidth selection, and a regularization term. The derivation of the optimal bandwidth can result in a decimal. In line with Cattaneo et al. (2019), we used the floor of the decimal. We increased the MSE optimal bandwidth by one, two, and three years respectively as a robustness check. The table displays the local average treatment effect for the birth cohort of 1976 and confidence intervals resulting from robust bias-corrected standard errors.*

## Footnotes

1 To our knowledge, no data on the TB related mortality rate were available. Thus, we used the absolute numbers of TB cases and divided them by the population size in the respective year (Statistics Norway; The World Bank (2022))

2 Giamarellos-Bourboulis et al. (2020) conducted a clinical trial with 202 participants aged 65 years and older who either received the BCG vaccine or a placebo at hospital discharge. The trial showed that the short-term risk of infection was reduced among the treatment group. However, these results cannot be generalized to a broader population nor long-term survival.

3 If there were multiple potential counterfactual countries, we could have combined them using a synthetic control approach. This was our initial intention after identifying seven potential control countries: Austria, Finland, France, Great Britain, Japan, Norway, and Portugal. Based on initial synthetic control analyses, we proceeded with the multi-period DiD rather than the synthetic control approach as no country other than Norway had a pre-trend that resembled Sweden, and as such the synthetic control approach gave nearly 100% of the weight to Norway.

